# On the road to early detection: A survey study of barriers and facilitators to community participation in a mobile lung cancer screening program

**DOI:** 10.64898/2026.04.15.26350954

**Authors:** Cherell Cottrell-Daniels, Najy Sadig, Sofia Haddan, Sashanna Roman, Vani N. Simmons, Matthew B. Schabath

## Abstract

**Background:** While a mobile lung cancer screening (mLCS) program can mitigate barriers to access, this study conducted a survey study to assess barriers and facilitators to mLCS which could inform the implementation of new mLCS programs or inform modifications to existing programs.

**Methods:** Patient eligibility included current age of 50 to 80 and had undergone any cancer screening at Moffitt Cancer Center (MCC) between January 1, 2023 and December 1, 2024. A web-based survey was administered from May 2025 to June 2025 which collected data on health behaviors, barriers, facilitators, screening preferences, and demographics. Descriptive statistics were used to quantify survey responses.

**Results:** Among participants who completed the survey, 73.4% reported no concerns about getting screened in a mobile screening unit, 67.9% reported concerned about the cost or if insurance covered mobile lung cancer screening, and 82.4% reported they would be screened if a voucher or insurance would pay for it. For preferences, 54.1% reported no preference for the time of year for a mobile screening event, 59.6% reported they will be willing to wait up to 30 minutes to get screened, and 44% would travel more than 20 minutes to get screened. There were no statistically significant differences in barriers and facilitators when the analyses were stratified by LCS eligibility.

**Conclusions:** We found acceptability of mobile lung cancer screening and preferences that are actionable including daytime weekday events, indoor waiting, short waits, proximity to home, clear cost coverage, and streamlined clinician recommendation.

## Introduction

Lung cancer remains the leading cause of cancer death in the United States, largely attributed to most patients diagnosed with late-stage disease [1]. The seminal lung cancer screening trial in the United States, the National Lung Screening Trial, demonstrated a more than 20% lung cancer–specific mortality reduction through low-dose computed tomography (LCDT) [2]. Based on this evidence, the National Comprehensive Cancer Network (NCCN) published their first set of lung cancer screening guidelines in 2012 [3] and the United States Preventive Services Task Force (USPSTF) issued lung cancer screening guidelines in 2013 [4]. Despite the availability of lung cancer screening for nearly 15 years, uptake of this life-saving modality has been dismal. Based on the 2021 USPSTF guidelines, in 2022 over 13 million Americans were eligible to get screened but uptake was only 16.4% [5]. Prior studies have identified several factors that contribute to low screening uptake including provider recommendations, limited access to healthcare services, fear of a cancer diagnosis, low perceived risk of lung cancer, stigmatization, cultural factors, and practical barriers of cost, insurance, transportation, and time off work [6-10].

Mobile screening units (MSUs) are an innovative alternative to fixed “brick and mortar” clinics. MSUs may include vans, recreational vehicles, or other traveling facilities staffed by health workers and outfitted with equipment for cancer screenings [11]. MSUs allow health care providers to increase their capacity outside of fixed clinics, which is particularly important in areas without an infrastructure for or access to cancer screening services MSUs increase community access by offering screening in convenient locations and times, thus decreasing the distance and travel time needed to access screening services. As an example for their utility, MSUs for breast cancer have indeed demonstrated greater reach among medically underserved populations [12]. Additionally, MSUs could further improve lung cancer outcomes, decrease downstream healthcare costs, and ultimately save lives. However, individuals with abnormal exams commonly need to report to a specialty clinic for follow-up.

A pivotal step in implementing an MSU is identifying potential barriers that may impact utilization. Therefore, the goal of this study was to conduct a survey study, using validated survey measures, to assess barriers and facilitators to mobile lung cancer screening (mLCS). Findings from this work can inform the implementation of new mLCS programs and guide modifications of existing mLCS programs, ultimately supporting broader uptake.

## Methods

### Study population

Participants were identified through the institutional Cancer Registry data and billing data. Eligibility included individuals who can read and write English, have a valid email address on record, current age of 50 to 80, and had undergone any cancer screening at Moffitt Cancer Center (MCC) between January 1, 2023 and December 1, 2024. Based on these inclusion criteria, we identified 5,345 patients. We focused on patients who underwent any cancer screening (i.e., lung, breast, colon, prostate, skin, thyroid, etc.,) since they have already engaged with preventative services and would have key insights with regards to facilitators, whereas those who meet lung cancer screening criteria may highlight unique barriers. This approach can help to optimize screening by understanding both perspectives. Additionally, among the 5,345 patients that were identified, 64.9% did not have complete smoking history data to determine LCS eligibility and only 7% had adequate smoking history data and were eligible for LCS (**Supplemental Figure 1**). As such, we could not prioritize patients who met LCS eligibility.

Using the institutional Cancer Registry, vital status was verified for all patients prior to initiating recruitment. Participants were recruited via email starting on May 21, 2025, followed by four email reminders sent approximately every five days. After the fourth reminder, a targeted follow-up email reminder was sent to participants who started the survey but did not complete it. The survey closed on June 25, 2025. A $10 Amazon gift card was offered to those who completed the survey. Individuals could opt-out and complete a survey that collected reasons for declining including, age, race, sex assigned at birth, and ethnicity. This study was approved by Advarra Inc. (Columbia, Maryland) Institutional Review Board.

### Survey

The survey was administered via REDCap and included items derived from validated measures [13, 14], height were collected to assess body mass index. Tobacco use was assessed by lifetime cigarette smoking, total years smoked, average number of cigarettes per day, and current smoking status. The items for weight, height, and tobacco use were taken from institutional questionnaire for lung cancer screening patients. **Barriers:** Factors that may affect participation in a mobile screening event as potential barriers. These items were developed by the team based on existing literature identifying barriers to engaging with mobile unit services more broadly [9, 15, 16]. Barriers included hours of operation of the mobile unit, confidentiality, perceived stigma, comfort level, trust in new healthcare providers, convenience, and accessibility. **Facilitators:** Factors that facilitate the uptake of mLCS were included, such as proximity to a mobile screening site or a hospital/clinic, receiving a recommendation from a healthcare provider, and their chosen/preferred sources for medical information [13, 14].

#### Screening preferences

These questions asked about the perceived accuracy of mobile screening, comfort with decision-making, modality preferences for a follow-up appointment, and the likelihood of participating if additional steps are required after the screening. These questions were developed using extant literature on participant uptake of mobile screening unit services [16]. **Open-ended questions:** Participants were asked to provide additional comments that may not be captured by quantitative measures. This included locations they would not like to be screened and any additional comments or thoughts about mobile lung cancer screening. **Demographic Information:** The survey collected demographics including age, race, ethnicity, sex, gender identity, years of education, military veteran status, marital status, employment status, and annual household income.

### Statistical Analysis

Descriptive statistics were used to quantify demographic characteristics and survey responses. For survey questions that had a “select all that apply” option, an “infrequent responses” category was created usually represented by responses at less than 1%; however, all individual responses are reported in the supplemental materials. Stratified analyses by LCS eligibility, age, race, and veteran status were conducted and differences were tested using Fisher’s Exact Chi-square test for categorical variables and Student’s *t* test for continuous variables. For the stratified analyses by LCS eligibility, we used 2026 NCCN guidelines criteria (≥ 50 years of age and 20 pack-years of smoking or ≥ 50 years of age and 20 years smoked). For the stratified analyses by age, we used 67 years (≥ 67 vs < 67) since this is the full retirement age for Social Security in the United States. All statistical tests were two-sided, and a *P*-value of less than .05 was considered statistically significant. All analyses for the quantitative data were conducted with Stata/MP version 14.2 (College Station, Texas).

For open-ended responses, a thematic analysis was conducted by the first author who has specialized training and expertise in qualitative research methods [17]. This approach ensured consistent and rigorous coding procedures and theme development throughout the analysis. To enhance credibility of the findings, the themes and interpretations were reviewed and discussed with the co-authors, allowing for additional perspectives and validation.

## Results

### Quantitative data

Of the 5,345 eligible patients that were emailed an invitation to participate, 218 participants completed the survey (**Table 1**), 137 started but did not complete the survey, and 23 declined to participate. Among the 218 who completed the survey, 169 participants requested an Amazon gift card. Among those who declined to participate, 8 completed the opt-out survey (**Supplemental Table 1**).

**Table 1.** Self-reported demographic characteristics.

| Characteristic | Overall<br>N = 218 | Not Eligible for LCS<br>(N=140) | Eligible for LCS<br>(N=78) | P-Value** |
| --- | --- | --- | --- | --- |
| **Age, mean (SD)\*** | 63.9 (8.1) | 62.9 (7.8) | 65.8 (8.2) | **0.013** |
| **Race (select all that apply), n (%)** |  |  |  | 0.402 |
| White or Caucasian | 195 (89.3) | 127 (90.8) | 68 (87.1) |  |
| Black or African American | 11 (5.0) | 8 (5.7) | 3 (3.8) |  |
| Asian | 3 (1.4) | 1 (0.7) | 2 (2.6) |  |
| Hawaiian or Pacific Islander | 0 (0) | 0 (0) | 0 (0) |  |
| American Indian or Alaska Native | 0 (0) | 0 (0) | 0 (0) |  |
| American Indian, Black, and White | 1 (0.5) | 1 (0.7) | 0 (0) |  |
| Black and Other | 1 (0.5) | 1 (0.7) | 0 (0) |  |
| Don't know/Prefer not to answer | 3 (1.4) | 1 (0.7) | 2 (2.6) |  |
| Something else/Not Sure (please specify): | 1 (0.5) | 0 (0) | 1 (1.3) |  |
| Missing | 3 (1.4) | 1 (0.7) | 2 (2.6) |  |
| **Hispanic, Latino, or Spanish origin (select all that apply), n (%)** |  |  |  | 0.214 |
| No not of Hispanic, Latino or Spanish origin | 189 (86.7) | 123 (87.9) | 66 (84.6) |  |
| Yes, Puerto Rican | 7 (3.2) | 5 (3.6) | 2 (2.6) |  |
| Yes, another Hispanic, Latino or Spanish origin (please specify): | 4 (1.8) | 3 (2.1) | 1 (1.3) |  |
| Yes, Cuban | 3 (1.4) | 0 (0) | 3 (3.8) |  |
| Yes, Mexican, Mexican American, Chicano/a | 3 (1.4) | 3 (2.1) | 0 (0) |  |
| Don't Know or Prefer Not to Answer | 3 (1.4) | 2 (1.4) | 1 (1.3) |  |
| Missing | 9 (4.1) | 4 (2.9) | 5 (6.4) |  |
| **Sex assigned at birth** |  |  |  | 0.113 |
| Female | 168 (77.1) | 112 (80.0) | 56 (71.8) |  |
| Male | 50 (22.9) | 28 (20.0) | 22 (28.2) |  |
| Don't Know or Prefer Not to Answer | 0 (0) | 0 (0) | 0 (0) |  |
| **Current gender identity** |  |  |  | 0.164 |
| Female | 165 (75.7) | 110 (78.6) | 55 (70.5) |  |
| Male | 48 (22.0) | 26 (18.6) | 22 (28.2) |  |
| I use a different term (please specify): | 1 (0.5) | 1 (0.7) | 0 (0) |  |
| Transgender | 0 (0) | 0 (0) | 0 (0) |  |
| Non-binary | 0 (0) | 0 (0) | 0 (0) |  |
| Two-spirit | 0 (0) | 0 (0) | 0 (0) |  |
| Don't know or prefer not to answer | 0 (0) | 0 (0) | 0 (0) |  |
| Missing | 4 (1.8) | 3 (2.1) | 1 (1.3) |  |
| **Highest level of education** |  |  |  | 0.169 |
| No formal schooling | 0 (0) | 0 (0) | 0 (0) |  |
| Less than 8th grade | 0 (0) | 0 (0) | 0 (0) |  |
| Some high school | 0 (0) | 0 (0) | 0 (0) |  |
| High school graduate/GED | 16 (7.3) | 10 (7.1) | 6 (7.7) |  |
| Technical/vocational training | 15 (6.9) | 8 (5.7) | 7 (9.0) |  |
| Associate degree/some college | 47 (21.6) | 24 (17.1) | 23 (29.5) |  |
| Bachelor's degree | 55 (25.2) | 39 (27.9) | 16 (20.5) |  |
| Post-graduate degree | 85 (39.0) | 59 (42.2) | 26 (33.3) |  |
| Do Not Know or Prefer Not to Answer | 0 (0) | 0 (0) | 0 (0) |  |

**Table 1. Continued**
| Characteristic | Overall<br>N = 218 | Not Eligible for LCS<br>(N=140) | Eligible for LCS<br>(N=78) | P-Value** |
| --- | --- | --- | --- | --- |
| **Have you smoked at least 100 cigarettes in your entire life?** |  |  |  | **< 0.001** |
| Yes | 132 (60.6) | 54 (38.6) | 78 (100.0) |  |
| No | 48 (22.0) | 48 (34.3) | 0 (0) |  |
| Never Smoked | 34 (15.6) | 34 (24.3) | 0 (0) |  |
| Don't know | 2 (0.9) | 2 (1.4) | 0 (0) |  |
| Prefer not to answer | 2 (0.9) | 2 (1.4) | 0 (0) |  |
| **Current smoker Smoke Cigarettes Now?** |  |  |  | **< 0.001** |
| Yes | 15 (6.9) | 2 (1.4) | 13 (16.7) |  |
| No | 163 (74.8) | 99 (70.7) | 64 (82.0) |  |
| Missing | 40 (18.3) | 39 (27.9) | 1 (1.3) |  |
| **Veteran status (select all that apply) – required question** |  |  |  | **0.300** |
| Not a veteran | 192 (88.1) | 125 (89.3) | 67 (85.9) |  |
| Retired or separated veteran | 17 (7.8) | 8 (5.7) | 9 (11.5) |  |
| Disabled veteran | 6 (2.7) | 4 (2.9) | 2 (2.6) |  |
| Retired or separated veteran, and Disabled veteran | 3 (1.4) | 3 (2.1) | 0 (0) |  |
| **Marital status** |  |  |  | **0.451** |
| Married | 146 (67.0) | 96 (68.6) | 50 (64.2) |  |
| Divorced | 33 (15.1) | 18 (12.9) | 15 (19.2) |  |
| Single | 18 (8.3) | 13 (9.3) | 5 (6.4) |  |
| Widowed | 12 (5.5) | 8 (5.7) | 4 (5.1) |  |
| Living with significant other | 6 (2.7) | 2 (1.4) | 4 (5.1) |  |
| Prefer not to answer | 2 (0.9) | 2 (1.4) | 0 (0) |  |
| Separated | 1 (0.5) | 1 (0.7) | 0 (0) |  |
| **Employment status** |  |  |  | **0.110** |
| Retired | 109 (50.0) | 66 (47.2) | 43 (55.1) |  |
| Employed | 79 (36.2) | 57 (40.7) | 22 (28.2) |  |
| Unable to work or disabled | 14 (6.4) | 8 (5.7) | 6 (7.7) |  |
| If other, please specify your employment | 9 (4.1) | 7 (5.0) | 2 (2.6) |  |
| Unemployed | 5 (2.3) | 1 (0.7) | 4 (5.1) |  |
| Student | 1 (0.5) | 1 (0.7) | 0 (0) |  |
| Don't know/Prefer not to answer | 0 (0) | 0 (0) | 0 (0) |  |
| Missing | 1 (0.5) | 0 (0) | 1 (1.3) |  |
| **Total family income per year** |  |  |  | **0.284** |
| \$60,000 or more per year | 123 (56.4) | 83 (59.4) | 40 (51.3) | |
| \$30,000 to \$60,000 per year | 30 (13.8) | 17 (12.1) | 13 (16.7) | |
| \$10,000 to \$30,000 per year | 16 (7.3) | 7 (5.0) | 9 (11.5) | |
| Less than \$10,000 per year | 2 (0.9) | 1 (0.7) | 1 (1.3) | |
| Don't know/Prefer not to answer | 46 (21.1) | 31 (22.1) | 15 (19.2) |  |
| Missing | 1 (0.5) | 1 (0.7) | 0 (0) |  |

**Table 1. Continued**
| Characteristic | Overall<br>N = 218 | Not Eligible for LCS<br>(N=140) | Eligible for LCS<br>(N=78) | P-Value** |
| --- | --- | --- | --- | --- |
| **Health care coverage -- required question** |  |  |  | **0.019** |
| Medicare | 86 (39.4) | 50 (35.7) | 36 (46.2) |  |
| Employer/union-provided health insurance (including plans purchased through another person's employer) | 85 (39.0) | 63 (45.1) | 22 (28.2) |  |
| A plan that you or another family member buys on your own | 15 (7.0) | 11 (7.9) | 4 (5.1) |  |
| TRICARE (formerly CHAMPUS), VA, or Military | 12 (5.5) | 8 (5.7) | 4 (5.1) |  |
| None (no coverage) | 7 (3.2) | 1 (0.7) | 6 (7.7) |  |
| Other (specify): | 7 (3.2) | 3 (2.1) | 4 (5.1) |  |
| Medicaid or other state program | 4 (1.8) | 2 (1.4) | 2 (2.6) |  |
| Indian Health Service | 0 (0) | 0 (0) | 0 (0) |  |
| Don't Know/Prefer not to answer | 2 (0.9) | 2 (1.4) | 0 (0) |  |

\*5 participants did not disclose their age
\*\***Bold** p-values are statistically significant

The mean age of study population who completed the survey (N = 218) was 63.9 years (SD 8.1). Most participants identified as White (89.3%), 5% identified as Black/African American, 1.4% as Asian, and 10% percent reported their ethnicity as Hispanic, Latino, or Spanish. Most respondents were assigned female at birth (77.1%) and 75.7% currently identified as female. For educational attainment, 39% completed postgraduate education and 25.2% a bachelor’s degree. Sixty-one percent self-reported as an ever smoker (i.e., ≥100 lifetime cigarettes), and 37.6% self-reported as a never smoker. Half were retired (50%), 36.2% were employed, and 56.4% reported annual household income ≥$60,000. The most common health coverage included Medicare (39.4%) and employer/union plans (39.0%). For the exploratory stratified analyses (**Supplemental Tables 2 – 4**), there were very few statistically significant differences for the demographic variables by age (**Supplemental Table 2**), race (**Supplemental Table 3**); and military service (**Supplemental Table 4**).

For the barriers and facilitators of community participation in mobile lung cancer screening (**Table 2**), the most frequently preferred screening time was weekdays 9am–5pm (44.5%), while 7.8% preferred evenings and weekends. Most respondents (73.4%) reported no concerns about being screened in a mobile unit. Among those who did express concerns, these were infrequent and included: accuracy (3.7%), privacy (2.8%), cleanliness (2.3%), equipment (1.4%), and temperature (0.9%). Being comfortable with a new (non-usual) clinician conducting the screening was very high (98.6%).

**Table 2.** Participant responses.

| Table 2. Participant responses |  |  |  |  |
| --- | --- | --- | --- | --- |
| Survey item | Overall<br>N = 218 | Not Eligible for LCS<br>(N=140) | Eligible for LCS<br>(N=78) | P-Value** |
| **Preferred days and times for screening if a mobile unit visited your neighborhood (select all that apply)** |  |  |  | 0.177 |
| ***Singular responses*** |  |  |  |  |
| M-F (9am-5pm) | 97 (44.5) | 57 (40.6) | 40 (51.3) |  |
| Mornings, M-F (before 9am) | 5 (2.3) | 5 (3.6) | 0 (0) |  |
| Evenings M-F (after 5pm) | 9 (4.1) | 4 (2.9) | 5 (6.4) |  |
| Weekends (9am-5pm) | 19 (8.7) | 17 (12.1) | 2 (2.6) |  |
| Weekends (before 9am) | 0 (0) | 0 (0) | 0 (0) |  |
| Weekends (after 5pm) | 1 (0.5) | 1 (0.7) | 0 (0) |  |
| ***Select all that apply responses*** |  |  |  |  |
| Evenings M-F (after 5pm) and weekends (9am-5pm) | 17 (7.8) | 11 (7.9) | 6 (7.7) |  |
| M-F (9am-5pm) and weekends (9am-5pm) | 16 (7.3) | 12 (8.6) | 4 (5.1) |  |
| All of the above | 8 (3.7) | 6 (4.3) | 2 (2.6) |  |
| M-F (9am-5pm), Evenings M-F (after 5pm), Weekends (9am-5pm), and Weekends (after 5pm) | 7 (3.2) | 4 (2.9) | 3 (3.8) |  |
| Infrequent Responses* | 39 (17.9) | 23 (16.4) | 16 (20.5) |  |
| **Concerns about getting screened in a mobile screening unit (select all that apply) -- required question** |  |  |  | 0.095 |
| ***Singular responses*** |  |  |  |  |
| No Concerns | 160 (73.4) | 101 (72.2) | 59 (75.7) |  |
| Accurate Results | 8 (3.7) | 5 (3.6) | 3 (3.8) |  |
| Privacy | 6 (2.8) | 6 (4.3) | 0 (0) |  |
| Cleanliness | 5 (2.3) | 2 (1.4) | 3 (3.8) |  |
| Equipment | 3 (1.4) | 0 (0) | 3 (3.8) |  |
| Temperature | 2 (0.9) | 2 (1.4) | 0 (0) |  |
| Other (Wheelchair accessibility) | 1 (0.5) | 1 (0.7) | 0 (0) |  |
| ***Select all that apply responses*** |  |  |  |  |
| Equipment, Cleanliness, and Accurate Results | 4 (1.8) | 2 (1.4) | 2 (2.6) |  |
| Equipment, Cleanliness, Temperature, Privacy, and Accurate Results | 4 (1.8) | 4 (2.9) | 0 (0) |  |
| Infrequent Responses* | 25 (11.4) | 17 (12.1) | 8 (10.3) |  |
| **Comfort with a new healthcare professional conducting the screening (for example, not associated with your primary care)? -- required question** |  |  |  | 0.700 |
| Yes | 215 (98.6) | 138 (98.6) | 77 (98.7) |  |
| No | 3 (1.4) | 2 (1.4) | 1 (1.3) |  |
| **Negative perceptions concerning getting screening services mobile units? (e.g., perceived as not being able to afford proper treatment)?** |  |  |  | 0.237 |
| Yes | 36 (16.5) | 24 (17.1) | 12 (15.4) |  |
| No | 137 (62.8) | 93 (66.5) | 44 (56.4) |  |
| Do Not Know or Prefer Not to Answer | 43 (19.8) | 23 (16.4) | 20 (25.6) |  |
| Missing | 2 (0.9) | 0 (0) | 2 (2.6) |  |
| **Concerned about the cost or if your insurance covered the mobile lung cancer screening?** |  |  |  | 0.329 |
| Yes | 148 (67.9) | 99 (70.7) | 49 (62.8) |  |
| No | 55 (25.2) | 33 (23.6) | 22 (28.2) |  |
| Do Not Know or Prefer Not to Answer | 14 (6.4) | 7 (5.0) | 7 (9.0) |  |
| Missing | 1 (0.5) | 1 (0.7) | 0 (0) |  |

| **Table 2. Continued** |  |  |  |  |
| --- | --- | --- | --- | --- |
| **Survey item** | **Overall**<br>**N = 218** | **Not Eligible for LCS**<br>**(N=140)** | **Eligible for LCS**<br>**(N=78)** | **P-**<br>**Value\*\*** |
| **Likely to attend a mobile lung cancer screening event in the summer?** |  |  |  | 0.388 |
| Very Likely | 82 (37.6) | 51 (36.5) | 31 (39.7) |  |
| Likely | 61 (28.0) | 35 (25.0) | 26 (33.3) |  |
| Neutral | 45 (20.6) | 33 (23.6) | 12 (15.4) |  |
| Unlikely | 23 (10.6) | 17 (12.1) | 6 (7.7) |  |
| Extremely Unlikely | 3 (1.4) | 2 (1.4) | 1 (1.3) |  |
| Missing | 4 (1.8) | 2 (1.4) | 2 (2.6) |  |
| **Good time of year for a mobile lung cancer screening event (select all that apply) -- required question** |  |  |  | 0.080 |
| ***Singular responses*** |  |  |  |  |
| No Preference | 118 (54.1) | 68 (48.7) | 50 (64.1) |  |
| Fall | 19 (8.7) | 15 (10.7) | 4 (5.1) |  |
| Winter | 17 (7.8) | 10 (7.1) | 7 (9.0) |  |
| Spring | 10 (4.6) | 5 (3.6) | 5 (6.4) |  |
| Summer | 8 (3.7) | 7 (5.0) | 1 (1.3) |  |
| ***Select all that apply responses*** |  |  |  |  |
| Spring, Fall, and Winter | 17 (7.8) | 13 (9.3) | 4 (5.1) |  |
| Fall and Winter | 8 (3.7) | 7 (5.0) | 1 (1.3) |  |
| Fall, Winter, Spring, and Summer | 6 (2.7) | 3 (2.1) | 3 (3.8) |  |
| Spring and Winter | 6 (2.7) | 6 (4.3) | 0 (0) |  |
| Spring and Fall | 3 (1.4) | 2 (1.4) | 1 (1.3) |  |
| Summer and Fall | 3 (1.4) | 3 (2.1) | 0 (0) |  |
| Infrequent Responses* | 3 (1.4) | 1 (0.7) | 2 (2.6) |  |
| **Length of time willing to wait for a screening – required question** |  |  |  | 0.151 |
| Less than 15 minutes | 48 (22.0) | 36 (25.7) | 12 (15.4) |  |
| 15-30 minutes | 130 (59.6) | 83 (59.3) | 47 (60.2) |  |
| 30-60 minutes | 35 (16.1) | 18 (12.9) | 17 (21.8) |  |
| More than 60 minutes | 5 (2.3) | 3 (2.1) | 2 (2.6) |  |
| **Where would you like the waiting room/area to be located?** |  |  |  | 0.293 |
| Indoor | 132 (60.6) | 91 (65.0) | 41 (52.6) |  |
| Outdoor | 10 (4.6) | 6 (4.3) | 4 (5.1) |  |
| No Preference | 48 (22.0) | 26 (18.6) | 22 (28.2) |  |
| Other | 26 (11.9) | 16 (11.4) | 10 (12.8) |  |
| Missing | 2 (0.9) | 1 (0.7) | 1 (1.3) |  |
| **Easiest location for mobile bus screening (select all that apply) – required question** |  |  |  | 0.705 |
| ***Singular responses*** |  |  |  |  |
| No preference | 75 (34.4) | 45 (32.1) | 30 (38.3) |  |
| Neighborhood | 16 (7.3) | 10 (7.1) | 6 (7.7) |  |
| Shopping mall/grocery store | 10 (4.6) | 7 (5.0) | 3 (3.8) |  |
| Downtown in my city | 4 (1.8) | 2 (1.4) | 2 (2.6) |  |
| Fire station or police station | 2 (0.9) | 2 (1.4) | 0 (0) |  |
| Church and other religious institutions | 1 (0.5) | 0 (0) | 1 (1.3) |  |
| School | 1 (0.5) | 1 (0.7) | 0 (0) |  |
| Health fair | 0 (0) | 0 (0) | 0 (0) |  |
| Post office | 0 (0) | 0 (0) | 0 (0) |  |
| Other | 2 (0.9) | 1 (0.7) | 1 (1.3) |  |
| **Survey item** | **Overall**<br>**N = 218** | **Not Eligible for LCS**<br>**(N=140)** | **Eligible for LCS**<br>**(N=78)** | **P-**<br>**Value\*\*** |
| **Select all that apply responses** |  |  |  |  |
| Neighborhood, and shopping mall/grocery store | 9 (4.1) | 7 (5.0) | 2 (2.6) |  |
| Neighborhood, shopping mall/grocery store, and Church or other religious institutions | 6 (2.8) | 4 (2.9) | 2 (2.6) |  |
| Neighborhood, Health Fair, and Church or other religious institutions | 4 (1.8) | 3 (2.1) | 1 (1.3) |  |
| Neighborhood, Fire station or police station, Church and other religious institutions | 4 (1.8) | 3 (2.1) | 1 (1.3) |  |
| Neighborhood, Fire station or police station, Shopping mall/grocery store | 4 (1.8) | 4 (2.9) | 0 (0) |  |
| Infrequent Responses* | 80 (36.8) | 51 (36.4) | 29 (37.2) |  |
| **Has your healthcare provider/doctor ever talked to you about getting screened for lung cancer?** |  |  |  | **<0.001** |
| Yes | 72 (33.0) | 23 (16.4) | 49 (62.8) |  |
| No | 126 (57.8) | 104 (74.3) | 22 (28.2) |  |
| Don't recall/ Don't remember | 20 (9.2) | 13 (9.3) | 7 (9.0) |  |
| **Previous experiences with cancer screenings? (select all that apply)** |  |  |  | **<0.001** |
| **Singular responses** |  |  |  |  |
| Breast | 15 (6.9) | 11 (7.9) | 4 (5.1) |  |
| Prostate | 5 (2.3) | 3 (2.1) | 2 (2.6) |  |
| Colon | 4 (1.8) | 2 (1.4) | 2 (2.6) |  |
| Lung | 4 (1.8) | 0 (0) | 4 (5.1) |  |
| Skin | 2 (0.9) | 1 (0.7) | 1 (1.3) |  |
| Cervical | 0 (0) | 0 (0) | 0 (0) |  |
| Other (GI, Skin and Thyroid, and bone marrow transplants) | 3 (1.4) | 1 (0.7) | 2 (2.6) |  |
| No previous experiences with cancer screenings | 3 (1.4) | 3 (2.1) | 0 (0) |  |
| **Select all that apply responses** |  |  |  |  |
| Breast, Colon, Skin, and Cervical | 35 (16.0) | 31 (22.1) | 4 (5.1) |  |
| Breast, Colon, Lung, Skin, and Cervical | 17 (7.8) | 9 (6.4) | 8 (10.3) |  |
| Breast, Colon, and Skin | 12 (5.5) | 10 (7.1) | 2 (2.6) |  |
| Colon, Prostate, and Skin | 11 (5.0) | 11 (7.9) | 0 (0) |  |
| Breast, Colon, and Cervical | 9 (4.1) | 8 (5.7) | 1 (1.3) |  |
| Breast, Skin, and Cervical | 8 (3.7) | 7 (5.0) | 1 (1.3) |  |
| Breast and Colon | 8 (3.7) | 5 (3.6) | 3 (3.8) |  |
| Infrequent Responses* | 79 (36.2) | 35 (25.2) | 44 (56.3) |  |
| Missing | 3 (1.4) | 3 (2.1) | 0 (0) |  |
| **Do you trust the health care system (for example, hospitals, pharmacies, and other organizations involved in health care)? -- required question** |  |  |  | **0.642** |
| Very | 120 (55.0) | 74 (52.9) | 46 (59.0) |  |
| Somewhat | 87 (40.0) | 58 (41.4) | 29 (37.2) |  |
| A little | 11 (5.0) | 8 (5.7) | 3 (3.8) |  |
| Not at all | 0 (0) | 0 (0) | 0 (0) |  |
| **Likely to get screened for lung cancer if you could get a voucher or insurance to pay for it?** |  |  |  | **0.647** |
| Extremely likely | 136 (62.4) | 87 (62.2) | 49 (62.8) |  |
| Likely | 48 (22.0) | 33 (23.6) | 15 (19.2) |  |
| Neutral | 27 (12.4) | 16 (11.4) | 11 (14.1) |  |
| Unlikely | 2 (0.9) | 1 (0.7) | 1 (1.3) |  |
| Extremely Unlikely | 3 (1.4) | 1 (0.7) | 2 (2.6) |  |
| Missing | 2 (0.9) | 2 (1.4) | 0 (0) |  |

Table 2. Continued
| Survey item | Overall<br>N = 218 | Not Eligible for LCS<br>(N=140) | Eligible for LCS<br>(N=78) | P-Value** |
| --- | --- | --- | --- | --- |
| **Would you be more likely to get screened for lung cancer if the screening could be done close to your home or in your neighborhood? -- required question** |  |  |  | 0.196 |
| Very likely | 126 (57.9) | 75 (53.5) | 51 (65.4) |  |
| Likely | 62 (28.4) | 46 (32.9) | 16 (20.5) |  |
| Neutral | 28 (12.8) | 18 (12.9) | 10 (12.8) |  |
| Unlikely | 0 (0) | 0 (0) | 0 (0) |  |
| Extremely Unlikely | 2 (0.9) | 1 (0.7) | 1 (1.3) |  |
| **Would you be more likely to be screened for lung cancer if your doctor/health care provider recommended it?** |  |  |  | 0.102 |
| Very Likely | 143 (65.6) | 91 (65.0) | 52 (66.6) |  |
| Likely | 48 (22.0) | 36 (25.7) | 12 (15.4) |  |
| Neutral | 24 (11.0) | 12 (8.6) | 12 (15.4) |  |
| Unlikely | 1 (0.5) | 0 (0) | 1 (1.3) |  |
| Extremely Unlikely | 2 (0.9) | 1 (0.7) | 1 (1.3) |  |
| **If you wanted more information about mobile lung cancer screening, how would you want to receive this information? (select all that apply):** |  |  |  | 0.279 |
| ***Singular responses*** |  |  |  |  |
| Receiving a notification in the mail | 26 (12.0) | 15 (10.7) | 11 (14.1) |  |
| Talking to your primary doctor | 20 (9.2) | 16 (11.4) | 4 (5.1) |  |
| Receiving a message through your electronic health record | 14 (6.4) | 5 (3.6) | 9 (11.5) |  |
| Receiving a phone call | 4 (1.8) | 3 (2.1) | 1 (1.3) |  |
| Through social media (for example, Facebook, Twitter, Instagram) | 2 (0.9) | 2 (1.4) | 0 (0) |  |
| Reading about this in a magazine/newspaper you subscribe to | 1 (0.5) | 1 (0.7) | 0 (0) |  |
| Watching a local news segment or television special about this special about this | 1 (0.5) | 0 (0) | 1 (1.3) |  |
| Other | 4 (1.8) | 1 (0.7) | 3 (3.8) |  |
| ***Select all that apply responses*** |  |  |  |  |
| Talking to your primary doctor, receiving a message through your electronic health record, and receiving a notification in the mail | 15 (6.8) | 10 (7.1) | 5 (6.4) |  |
| Talking to your primary doctor, and receiving a message through your electronic health record | 9 (4.1) | 8 (5.7) | 1 (1.3) |  |
| Receiving a message through your electronic health record, and receiving a notification in the mail | 8 (3.7) | 2 (1.4) | 6 (7.7) |  |
| Talking to your primary doctor, and receiving notification in the mail | 7 (3.2) | 6 (4.3) | 1 (1.3) |  |
| Talking to your primary doctor, receiving a phone call, receiving a message through your electronic health record, and receiving a notification in the mail | 7 (3.2) | 5 (3.6) | 2 (2.6) |  |
| Receiving a phone call, receiving a message through your electronic health record, and receiving a notification in the mail | 6 (2.8) | 4 (2.9) | 2 (2.6) |  |
| Talking to your primary doctor, Receiving a phone call, and Receiving a message through your electronic health record | 6 (2.8) | 4 (2.9) | 2 (2.6) |  |
| Infrequent Responses* | 86 (39.4) | 57 (40.8) | 29 (37.1) |  |
| Missing | 2 (0.9) | 1 (0.7) | 1 (1.3) |  |

| **Table 2. Continued** |  |  |  |  |
| --- | --- | --- | --- | --- |
| **Survey item** | **Overall**<br>**N = 218** | **Not Eligible for LCS**<br>**(N=140)** | **Eligible for LCS**<br>**(N=78)** | **P-**<br>**Value\*\*** |
| **Furthest distance you would travel to get screened (mobile unit or building-hospital/clinic)? -**<br>**- required question** |  |  |  | 0.343 |
| Less than 5 minutes | 0 (0) | 0 (0) | 0 (0) |  |
| 5-10 minutes | 24 (11.0) | 15 (10.7) | 9 (11.5) |  |
| 10-20 minutes | 98 (45.0) | 68 (48.6) | 30 (38.5) |  |
| More than 20 minutes | 96 (44.0) | 57 (40.7) | 39 (50.0) |  |
| **Would you be comfortable talking with your doctor about lung cancer screening in a virtual**<br>**appointment?** |  |  |  | 0.925 |
| Very comfortable | 151 (69.2) | 97 (69.3) | 54 (69.2) |  |
| Somewhat comfortable | 29 (13.3) | 19 (13.6) | 10 (12.8) |  |
| Neutral | 25 (11.5) | 17 (12.1) | 8 (10.3) |  |
| Somewhat uncomfortable | 1 (0.5) | 1 (0.7) | 0 (0) |  |
| Very uncomfortable | 11 (5.0) | 6 (4.3) | 5 (6.4) |  |
| Missing | 1 (0.5) | 0 (0) | 1 (1.3) |  |
| **If you needed a follow-up appointment after your lung cancer screening, where would you be**<br>**willing to have it? (select all that apply)** |  |  |  | 0.217 |
| ***Singular responses*** |  |  |  |  |
| Hospital/clinic Only | 49 (22.4) | 33 (23.6) | 16 (20.5) |  |
| Virtual Only | 2 (0.9) | 2 (1.4) | 0 (0) |  |
| Mobile Screening Unit Only | 0 (0) | 0 (0) | 0 (0) |  |
| Other (At my local healthcare office) | 5 (2.0) | 4 (2.9) | 1 (1.3) |  |
| Other (Moffitt) | 3 (1.2) | 1 (0.7) | 2 (2.6) |  |
| Other (Depends) | 1 (0.4) | 0 (0) | 1 (1.3) |  |
| ***Select all that apply responses*** |  |  |  |  |
| Hospital/Clinic, and Mobile Screening Unit | 12 (6.0) | 5 (3.6) | 7 (9.0) |  |
| Hospital/Clinic, Mobile Screening Unit, and Virtual | 132 (61.0) | 89 (63.5) | 43 (55.0) |  |
| Hospital/Clinic and Virtual | 9 (4.1) | 4 (2.9) | 5 (6.4) |  |
| Mobile Screening Unit, and Other | 1 (0.4) | 0 (0) | 1 (1.3) |  |
| Virtual and Other | 1 (0.4) | 1 (0.7) | 0 (0) |  |
| Hospital/clinic, Virtual, and Other | 1 (0.4) | 0 (0) | 1 (1.3) |  |
| All the above, and Other | 1 (0.4) | 0 (0) | 1 (1.3) |  |
| Hospital/Clinic, Mobile Screening Unit and Other | 1 (0.4) | 1 (0.7) | 0 (0) |  |

\*For survey questions that had a “select all that apply” option, an “Infrequent Responses” category was created. All individual responses are reported in the supplemental materials.
\*\***Bold** p-values are statistically significant

When asked about negative perceptions (e.g., stigma, inability to afford care, etc.,), 62.8% reported “no,” 16.5% “yes,” and 19.8% “don’t know/prefer not to answer”. Concerns about cost/coverage were common (67.9% reported “yes”). However, most indicated they would be more likely to get screened if a voucher or insurance covered the cost (62.4% extremely likely and 22% likely).

Regarding waiting time expectations, 22.0% are willing to wait <15 minutes, 59.6% 15–30 minutes, and 18.4% ≥30 minutes. An indoor waiting area was preferred (60.6%). Proximity was also important: 57.9% reported being “very likely” and 28.4% “likely” to screen if the service was available close to home. Provider recommendation was influential (65.6% “very likely,” 22.0% “likely” to screen if recommended). However, only 33.0% reported a prior clinician discussion about lung cancer screening.

The locale of the mobile screening unit, 34.4% reported “no preference”, with smaller proportions selecting neighborhood, shopping centers, or other specific locations. For willingness for time to travel to the mobile unit, 45% would travel 10–20 minutes and 44% >20 minutes. Most respondents were “very comfortable” (69.25) with virtual discussions about screening. For follow-up after screening, multiple care venues were acceptable: 61.0% selected hospital/clinic + mobile unit + virtual options; 22.4% preferred hospital/clinic only.

For stratified analyses of the barriers and facilitators, the only statistically significant differences by age (< 67 vs. ≥ 67**; Supplemental Table 5**) was willingness to get screened if covered by voucher or insurance and follow-up appointment location. The only statistically significant differences by race (Non-Hispanic White vs. everyone else) were concerns about cost/coverage, good time of year for screening event, and previous cancer screening experiences (**Supplemental Table 6**). The statistically significant differences by military service (veterans vs. non-veterans) were previous cancer screening experiences, willingness to get screened if covered by voucher or insurance, and comfortable with virtual discussions about screening (**Supplemental Table 7**).

**Supplemental Table 8** lists all responses for the “select all that apply” questions including infrequent responses.

#### Open-ended responses

Analysis of open-ended responses revealed several recurring themes. When asked how concerns about mobile screening could be addressed, participants most frequently emphasized ***equipment reliability and accuracy*** (n=38), followed by ***sanitization and cleanliness*** (n=32). ***Comfort related to temperature*** was also a notable issue (n=22), along with ***privacy during screening*** (n=20) and ***staff professionalism*** (n=12). Representative comments included requests for “proof of servicing equipment” and assurances that “all equipment [is] sanitized” between patients, highlighting trust and hygiene as critical factors (Table 3).

**Table 3.**
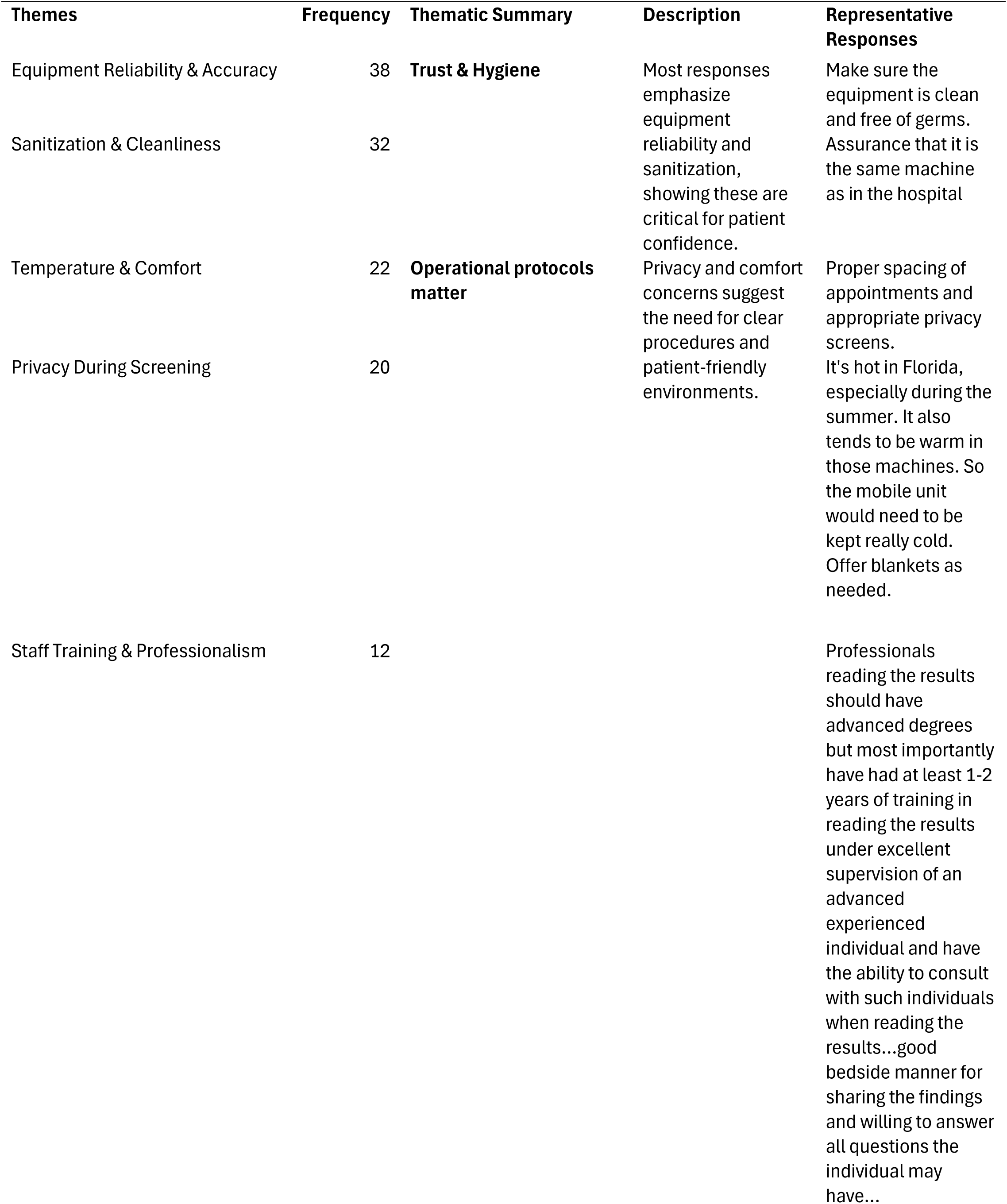
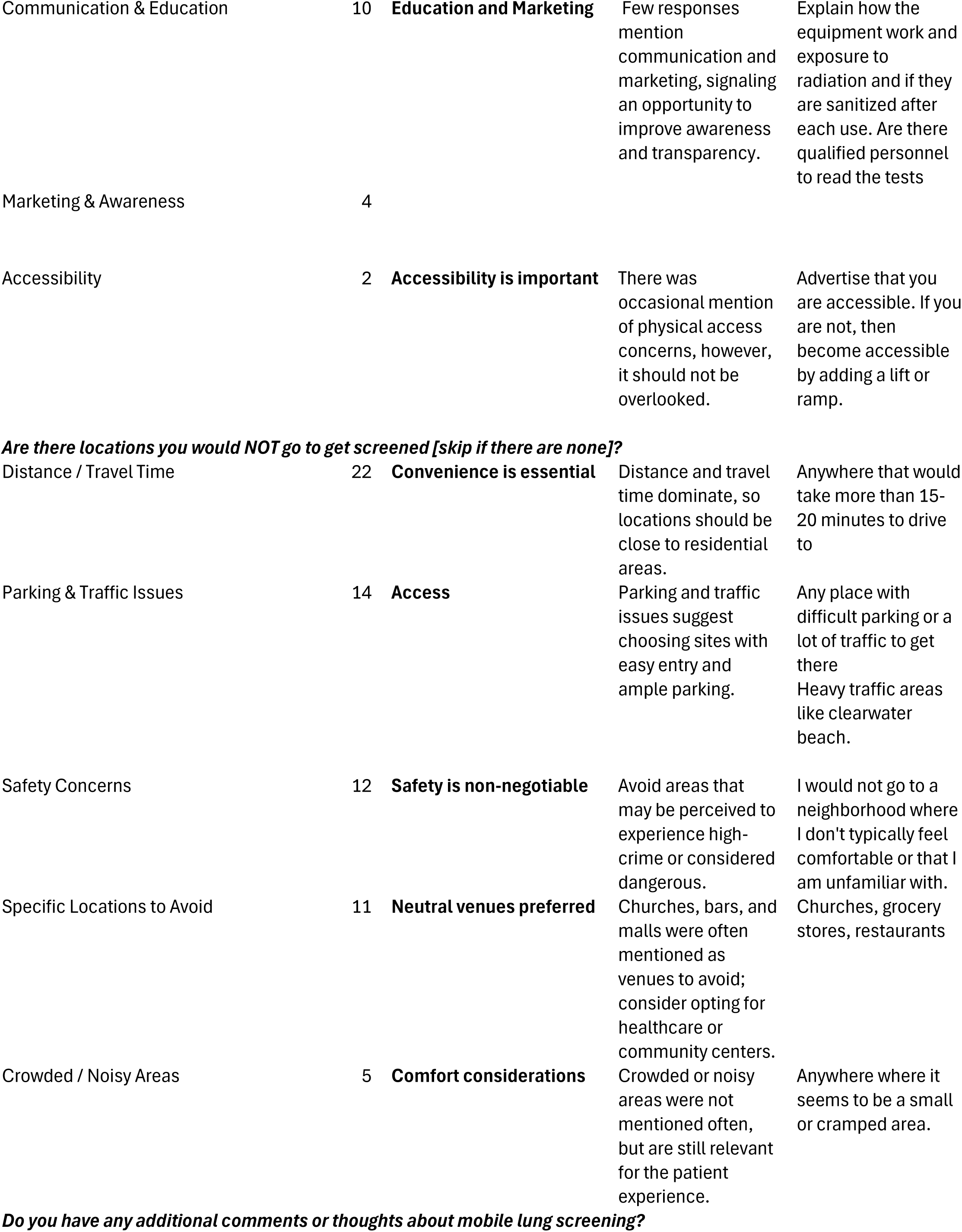
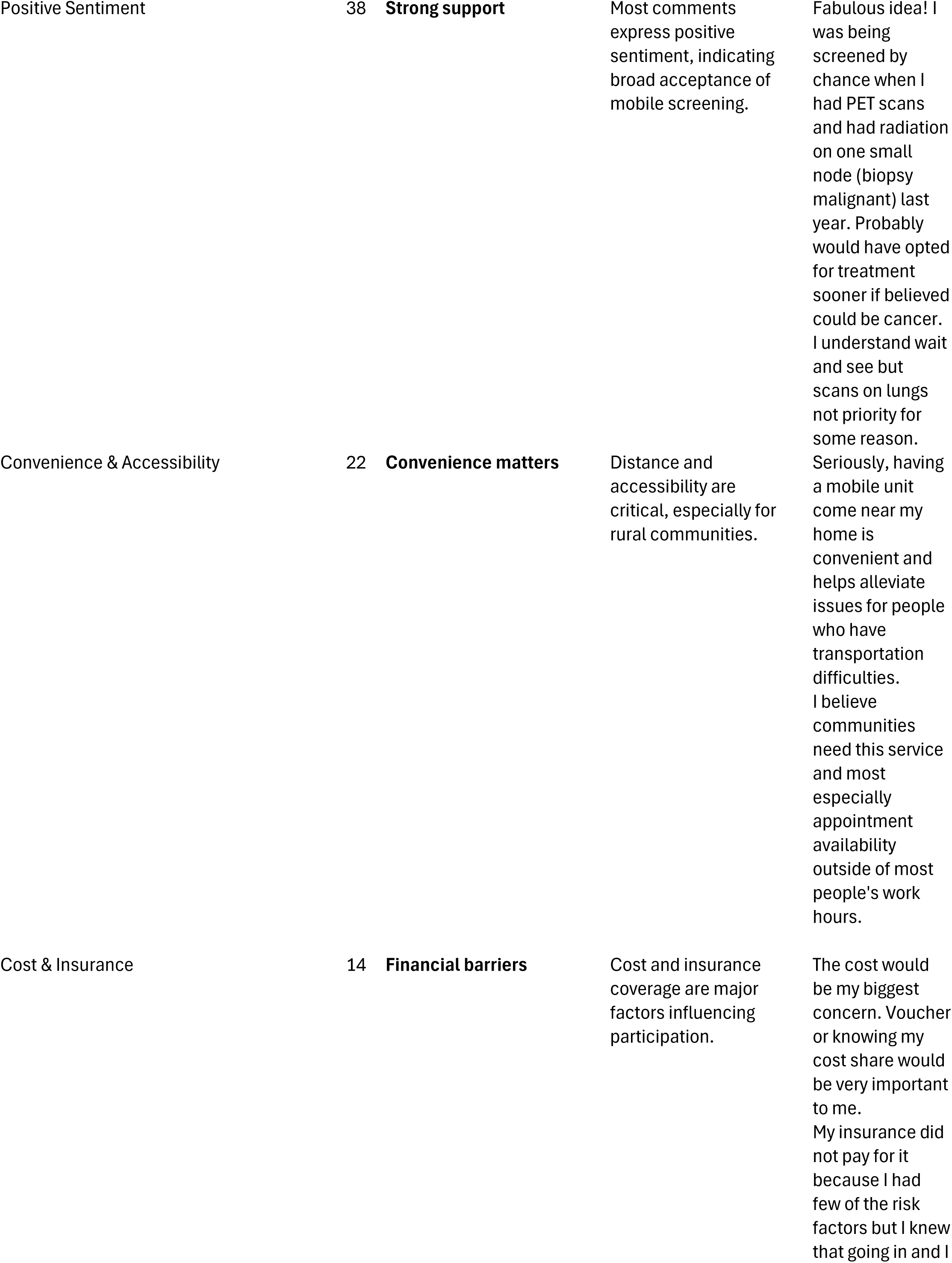
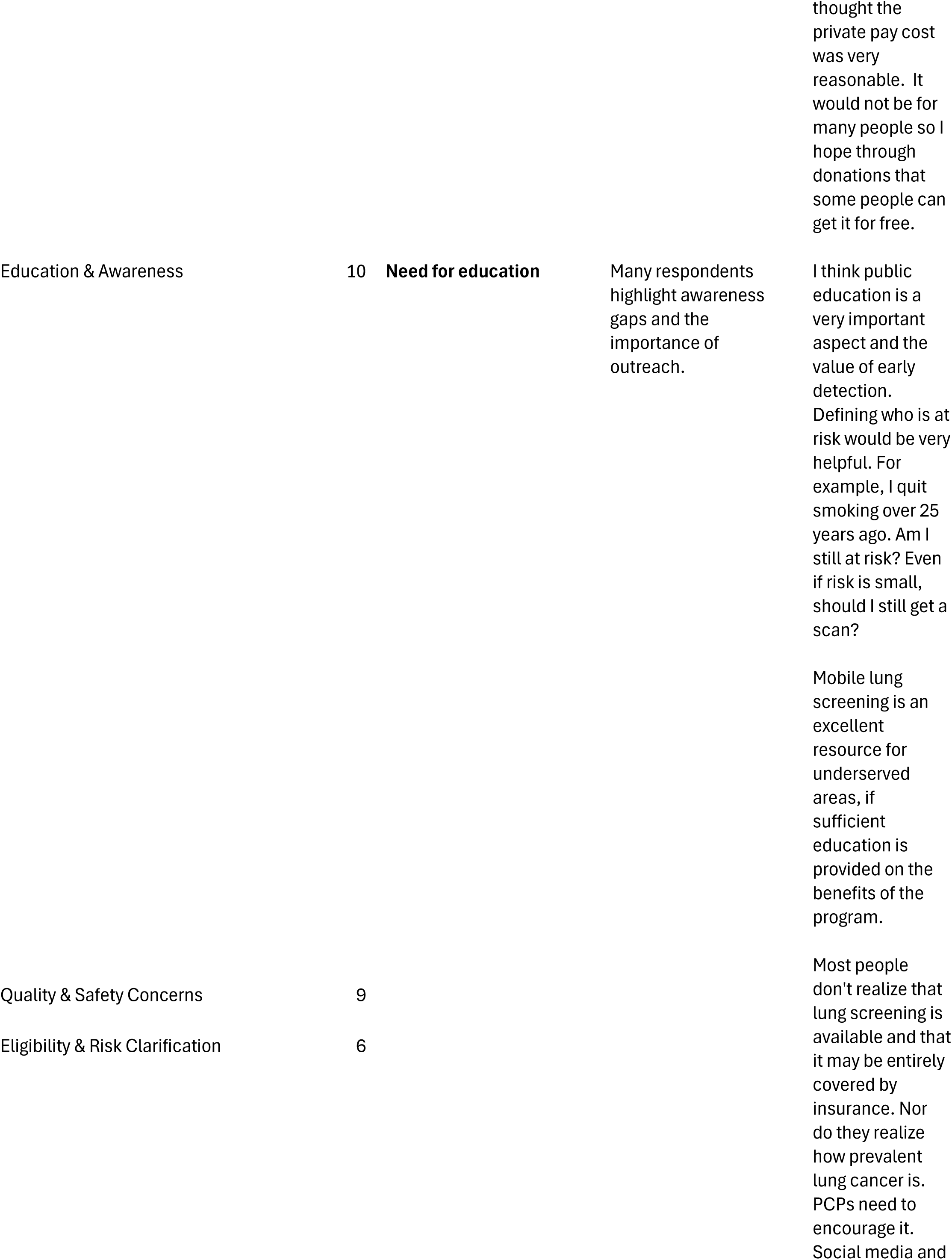
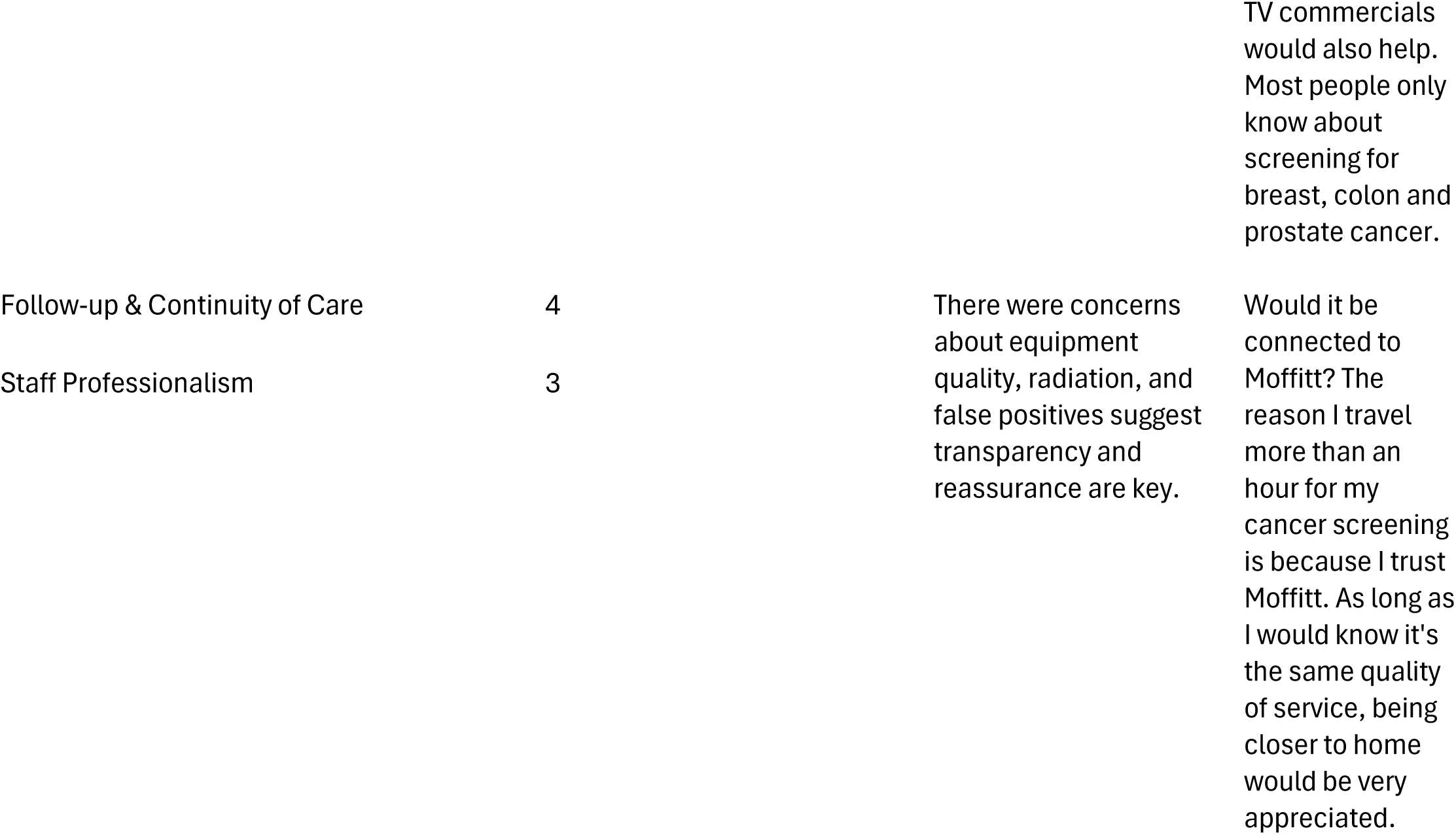
Summary of participants concerns and suggested strategies for a mobile lung cancer screening program. What can be done to address [participant response] concerns?

Regarding locations respondents would avoid, ***distance and travel time*** emerged as the common barrier (n=22), with many unwilling to travel more than 15–20 minutes. ***Parking and traffic issues*** (n=14) and ***safety concerns*** (n=12) were also common, with participants citing “sketchy neighborhoods” and “downtown areas with limited parking” as undesirable. Neutral venues such as healthcare or community centers were preferred over churches, bars, or malls (Table 3).

Finally, additional comments reflected strong ***positive sentiment*** toward mobile screening (n=38), with convenience and accessibility frequently mentioned (n=22). However, respondents also raised concerns about ***cost and insurance coverage*** (n=14) and called for greater ***education and awareness*** (n=10). Many expressed trust in established institutions, noting, “Would it be connected to Moffitt? … I trust Moffitt,” and emphasized the importance of clear eligibility criteria and follow-up care (**Table 3**).

## Discussion

In this survey study of a patient population derived from an NCI-designated Comprehensive Cancer Center, we found high acceptability of mobile lung cancer screening, strong responsiveness to clinician recommendation and cost coverage, and practical preferences that can guide implementation. Most participants expressed no concerns about mobile screening, were comfortable with a new clinician, and preferred indoor waiting and weekday daytime operations. Key facilitators to screening uptake included proximity to home, voucher/insurance coverage, and provider recommendation. Despite the importance of clinician influence, only one-third reported that a clinician had ever discussed lung cancer screening——highlighting an actionable gap in provider–patient communication that could be addressed to improve uptake. Importantly, while our study population included patients who underwent any cancer screening, we found no statistically significant differences in barriers and facilitators when we stratified the analyses by LCS eligibility.

The quantitative data revealed that a large majority of participants (73.4%) reported no concerns about mobile lung cancer screening, indicating broad acceptability. However, the thematic analysis provided important details by identifying specific concerns among those who did express reservations, particularly around equipment reliability, accuracy, and cleanliness. These qualitative insights highlight the critical role of trust in technology and hygiene practices, which may not be fully captured by quantitative survey questions but are essential for patient confidence and uptake.

The finding that only one-third of participants had discussed lung cancer screening with a clinician reflects a widespread implementation challenge documented in a recent systematic review [16]. Research demonstrates that provider awareness of screening guidelines significantly impacts utilization rates, with studies showing screening ordering rates of 71% versus 38% when providers are knowledgeable about USPSTF guidelines [18]. Recent evidence indicates that electronic health record alerts can dramatically improve screening uptake from 2.2% to 21.1% [19], while multifaceted interventions combining clinician-facing reminders, electronic health record-integrated shared decision-making tools, and narrative guidance (i.e., conversations from a provider to enhance the patient and provider experiences) can increase care gap closure from 15.9% to 46.9% [20]. For instance, a provider might share a personal story to help a patient cope with receiving negative test results. The robust effect of provider recommendations found in our study aligns with evidence showing that Medicare beneficiaries who participated in shared decision-making visits prior to initial screening had subsequent adherence rates that were 26.5-32.5% higher over four-year follow-up periods compared to those who did not participate in the shared-decision making visits [21].

The high acceptability of mobile care and low reported stigma in our sample challenge common assumptions that mobile delivery is inherently viewed as "second-class" care. A study of a mobile health clinic serving a vulnerable population (people who use drugs) demonstrated that clients perceived mobile units as beneficial to their communities, providing accessible care without stigma [22]. This positive perception is consistent with broader evidence showing that when mobile health services reduce access barriers without compromising care quality, they are viewed favorably by users. The acceptance of mobile screening found in our study supports mobile programs as viable alternatives to fixed-site facilities, particularly for populations facing geographic or logistical barriers to screening access.

Participants’ willingness to travel >20 minutes for screening supports regional scheduling approaches that align with evidence about geographic access barriers. The qualitative findings add depth by revealing that participants prefer screening sites close to home, in safe neighborhoods with ample parking, and neutral venues such as healthcare or community centers rather than commercial or religious spaces. A recent study found that approximately 5% of the eligible population lacked access to screening facilities within 40 miles [23]. Geographic analysis in the U.S. indicated that more than one-third of counties with high lung cancer mortality rates are beyond 60 minutes’ drive-time from screening centers, affecting approximately 60.5 million people or 19% of the country’s population [24]. Mobile screening units have been specifically identified as effective strategies to reach underserved individuals in their communities and reduce geographic disparities in access to lung cancer screening [8].

The strong stated effect of voucher/insurance coverage on acceptability reflects broader evidence about financial barriers to screening participation. The thematic analysis further elucidated this with participants’ discussing desire for clear communication about coverage, out-of-pocket costs, and eligibility criteria, as well as calls for increased education and awareness to reduce financial and informational barriers. A recent study on cervical cancer screening conducted among low-income, uninsured or publicly insured women aged 25-64 years, found that perceived financial barriers affect screening decisions across diverse populations, with screening appointment costs and follow-up treatment expenses being primary concerns [25]. A recent systematic review on patient barriers and facilitators for lung cancer screening uptake demonstrated that cost transparency and coverage assurance are critical implementation factors, with financial cost being the most common barrier to screening uptake [10]. Our findings reinforce the importance of explicit messaging about no-cost screening and streamlined financial counseling in mLCS program design.

The willingness to engage in virtual communication and acceptance of multiple follow-up venues found in our study support hybrid models that combine remote pre-visit counseling, mobile unit imaging, and flexible follow-up options [26]. This aligns with emerging evidence on hybrid telehealth approaches that combine in-person and remote care modalities to reduce travel burdens while maintaining care quality. Research shows that hybrid models can enable continuous management, improve care access, and provide flexible service delivery without compromising quality outcomes [27, 28]. The integration of virtual components with mobile screening represents an innovative approach to addressing multiple barriers simultaneously.

While not directly assessed in our study, the trust in provider recommendations we identified supports integration of patient navigation roles within mLCS programs. Recent systematic reviews demonstrated that patient navigation programs can improve screening rates, compliance with follow-up, and time to treatment initiation, particularly among vulnerable populations [29, 30]. Recent studies show patient navigation significantly improves lung cancer screening completion, with particularly strong effects among populations facing housing instability [31] and other social barriers [32]. The implementation of navigation services within mLCS programs could address both logistical barriers and the communication gaps identified in our findings.

Our findings about the importance of provider recommendations support the integration of evidence-based shared decision-making tools within mLCS programs. One systematic review identified 15 distinct tools to support shared decision-making for lung cancer screening, with studies showing improvements in patient knowledge and reductions in decisional conflict [33]. While the effectiveness of these tools varies, research demonstrates that decision aids can improve patient knowledge and support informed decision-making when integrated into clinical workflows [34]. The integration of such tools within mLCS programs could enhance the quality of provider-patient discussions while ensuring compliance with Centers for Medicare & Medicaid Services requirements.

The key findings provide guidance for comprehensive mLCS program design that addresses documented barriers at multiple levels. First, the predominance of weekday daytime preferences, combined with short acceptable wait times, suggests integrating mLCS events with community hubs during peak accessibility hours. Second, the combination of cost concerns and strong voucher effects argues for comprehensive financial navigation and benefits verification. Third, the provider recommendation affects support systematic approaches, including electronic health record prompts, scripted counseling protocols, and referral workflows to address documented communication gaps. Fourth, the acceptance of hybrid approaches supports models combining remote counseling, mobile imaging, and flexible follow-up that can reduce friction while maintaining care quality.

Strengths of this study include its use of a diverse, comprehensive cancer center patient population, validated survey items addressing multilevel barriers and facilitators, and a practical focus on operational preferences and cost considerations. While there are several strengths, the current study is not without limitations. The study used email recruitment from a single tertiary cancer center, with a low response proportion relative to invitations (218 out of 5,345), raising potential selection bias. The sample was largely White and highly educated, which may limit generalizability to communities prioritized for mLCS equity initiatives.

## Conclusions

In this study, mLCS was deemed broadly acceptable by very high comfort and low concerns, with preferences that are actionable including: daytime weekday events, indoor waiting, short waits, proximity to home, clear cost coverage, and streamlined clinician recommendation. Implementation strategies that center on provider prompts/referrals, financial transparency (vouchers/coverage), and hybrid workflows may enhance uptake.

## Acknowledgement

This work has been supported in part by the Collaborative Data Services Core (CDSC) and Participant Research, Interventions, and Measurements (PRISM) Core at the H. Lee Moffitt Cancer Center & Research Institute, an NCI designated Comprehensive Cancer Center (P30 CA076292).

## Data Availability

The data will be made available upon reasonable request.

## Supplemental Materials

### Supplemental Figure Legends

**Supplemental Figure 1.**
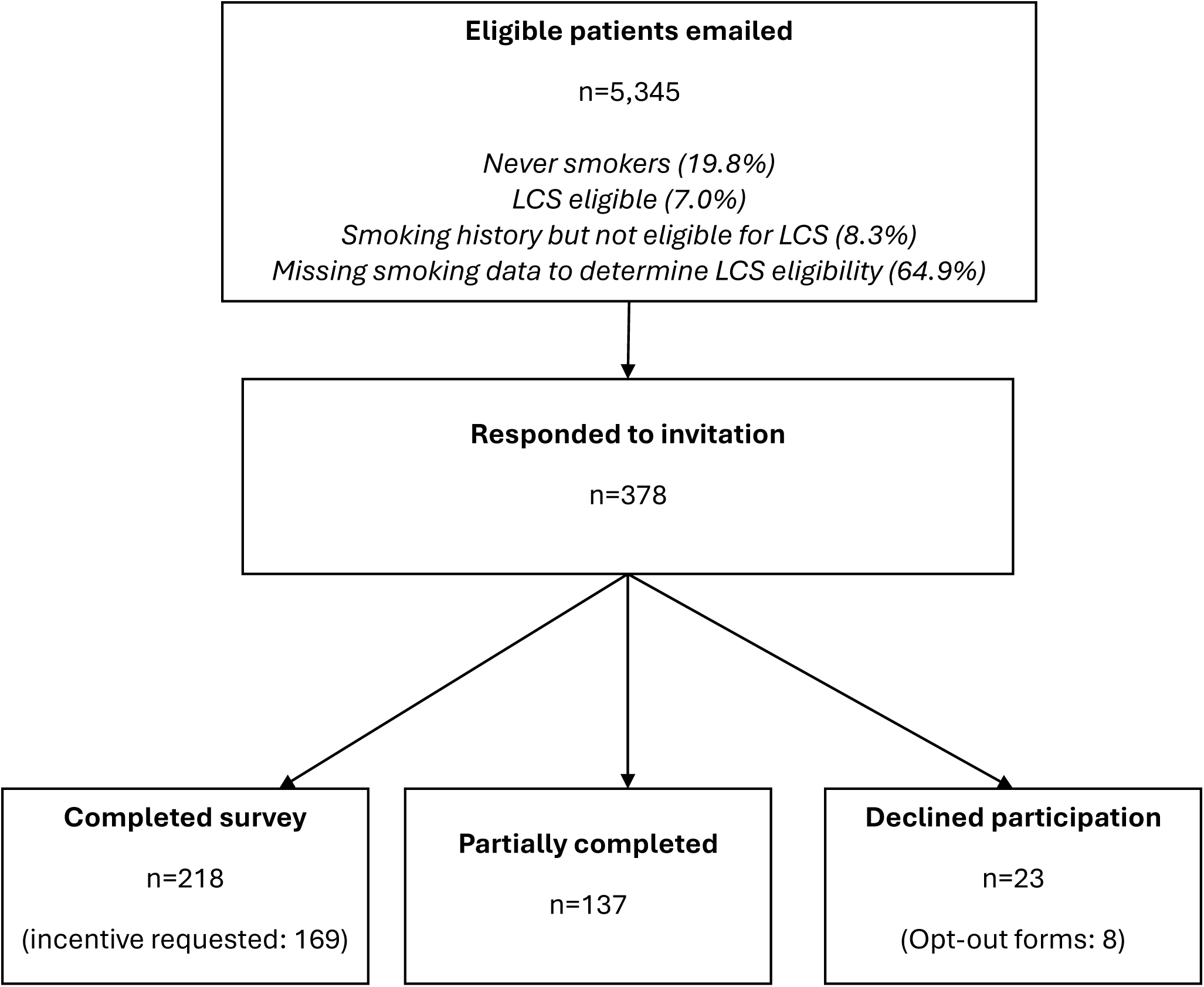
CONSORT diagram for recruitment of participants

**Supplemental Table 1.** Opt-outs.

| **Supplemental Table 1. Opt-outs** |  | **N=8\*** |
| --- | --- | --- |
| **Age: Mean (SD)** |  | 65.1 (SD 8.7) |
| **Race (select all that apply)** |  |  |
| White |  | 6 (75.0) |
| Asian |  | 0 (0) |
| Black or African American |  | 0 (0) |
| American Indian or Alaskan Native |  | 0 (0) |
| Native Hawaiian or other Pacific Islander |  | 0 (0) |
| Multi-racial |  | 0 (0) |
| Other |  | 0 (0) |
| Prefer not to answer |  | 1 (12.5) |
| Missing |  | 1 (12.5) |
| **Hispanic, Latino, or Spanish origin (select all that apply)** |  |  |
| Puerto Rican |  | 1 (12.5) |
| Cuban |  | 0 (0) |
| Dominican Republic |  | 0 (0) |
| Spanish |  | 0 (0) |
| Mexican, Mexican American, or Chicano |  | 0 (0) |
| Central of South American, other than Brazilian |  | 1 (12.5) |
| Brazilian |  | 0 (0) |
| Other |  | 0 (0) |
| Prefer not to answer |  | 0 (0) |
| None-Not Hispanic, Latino, or Latin X |  | 5 (62.5) |
| Missing |  | 1 (12.5) |
| **Sex assigned at birth** |  |  |
| Female |  | 3 (37.5) |
| Male |  | 4 (50.0) |
| Don't Know or Prefer Not to Answer |  | 0 (0) |
| Missing |  | 1 (12.5) |

\*Missing responses were not from the same individual

**Supplemental Table 2.** Demographics by age.

| **Supplemental Table 2. Demographics by age** |  |  |  |
| --- | --- | --- | --- |
| **Characteristic** | **< 67**<br>**(N=130)** | **≥ 67**<br>**(N=83)** | **P-value** |
| **Race** |  |  | **0.111** |
| White or Caucasian | 116 (89.2) | 75 (90.4) |  |
| Black or African American | 8 (6.2) | 3 (3.6) |  |
| Asian | 0 (0) | 2 (2.4) |  |
| Hawaiian or Pacific Islander | 0 (0) | 0 (0) |  |
| American Indian or Alaska Native | 0 (0) | 0 (0) |  |
| American Indian, Black, and White | 0 (0) | 1 (1.2) |  |
| Black and Other | 1 (0.8) | 0 (0) |  |
| Don't know/Prefer not to answer | 3 (2.3) | 0 (0) |  |
| Something else/Not Sure (please specify): | 0 (0) | 1 (1.2) |  |
| Missing | 2 (1.5) | 1 (1.2) |  |
| **Hispanic, Latino, or Spanish origin** |  |  | **0.257** |
| No not of Hispanic, Latino or Spanish origin | 111 (85.4) | 75 (90.3) |  |
| Yes, Puerto Rican | 7 (5.4) | 0 (0) |  |
| Yes, another Hispanic, Latino or Spanish origin<br>(please specify): | 3 (2.3) | 1 (1.2) |  |
| Yes, Cuban | 1 (0.8) | 1 (1.2) |  |
| Yes, Mexican, Mexican American, Chicano/a | 2 (1.5) | 1 (1.2) |  |
| Don't Know or Prefer Not to Answer | 1 (0.8) | 1 (1.2) |  |
| Missing | 5 (3.8) | 4 (4.9) |  |
| **Sex assigned at birth** |  |  | **0.020** |
| Female | 107 (82.3) | 56 (67.5) |  |
| Male | 23 (17.7) | 27 (32.5) |  |
| Don't Know or Prefer Not to Answer | 0 (0) | 0 (0) |  |
| **Current gender identity** |  |  | **0.022** |
| Female | 105 (80.8) | 55 (66.3) |  |
| Male | 23 (17.7) | 25 (30.1) |  |
| I use a different term (please specify): | 0 (0) | 1 (1.2) |  |
| Transgender | 0 (0) | 0 (0) |  |
| Non-binary | 0 (0) | 0 (0) |  |
| Two-spirit | 0 (0) | 0 (0) |  |
| Don't know or prefer not to answer | 0 (0) | 0 (0) |  |
| Missing | 2 (1.5) | 2 (2.4) |  |
| **Highest level of education** |  |  | **0.168** |
| No formal schooling | 0 (0) | 0 (0) |  |
| Less than 8th grade | 0 (0) | 0 (0) |  |
| Some high school | 0 (0) | 0 (0) |  |
| High school graduate/GED | 12 (9.2) | 4 (4.8) |  |
| Technical/vocational training | 12 (9.2) | 3 (3.6) |  |
| Associate degree/some college | 28 (21.5) | 18 (21.7) |  |
| Bachelor's degree | 34 (26.3) | 18 (21.7) |  |
| Post-graduate degree | 44 (33.8) | 40 (48.2) |  |
| Do Not Know or Prefer Not to Answer | 0 (0) | 0 (0) |  |
| **Have you smoked at least 100 cigarettes in your entire life?** |  |  | **0.546** |
| Yes | 77 (59.2) | 51 (61.4) |  |
| No | 26 (20.0) | 21 (25.4) |  |
| Never smoked | 24 (18.5) | 10 (12.0) |  |
| Don't know | 2 (1.5) | 0 (0) |  |
| Prefer not to answer | 1 (0.8) | 1 (1.2) |  |

| **Supplemental Table 2. Continued** |  |  |  |
| --- | --- | --- | --- |
| **Characteristic** | **< 67**<br>**(N=130)** | **≥ 67**<br>**(N=83)** | **P-value** |
| **Current Smoker** |  |  | 0.102 |
| No | 90 (69.2) | 69 (83.1) |  |
| Yes | 12 (9.2) | 3 (3.6) |  |
| Missing | 28 (21.6) | 11 (13.3) |  |
| **Veteran Status** |  |  | 0.285 |
| I am NOT a veteran | 118 (90.8) | 69 (83.2) |  |
| Retired or separated veteran | 7 (5.4) | 10 (12.0) |  |
| Disabled veteran | 3 (2.3) | 3 (3.6) |  |
| Retired or separated veteran, and Disabled veteran | 2 (1.5) | 1 (1.2) |  |
| **Marital Status** |  |  | 0.213 |
| Married | 90 (69.2) | 53 (63.9) |  |
| Divorced | 19 (14.6) | 13 (15.7) |  |
| Single | 12 (9.3) | 6 (7.2) |  |
| Widowed | 5 (3.8) | 6 (7.2) |  |
| Living with significant other | 1 (0.8) | 5 (6.0) |  |
| Prefer not to answer | 2 (1.5) | 0 (0) |  |
| Separated | 1 (0.8) | 0 (0) |  |
| **Employment Status** |  |  | <0.001 |
| Employed | 66 (50.8) | 13 (15.7) |  |
| Retired | 41 (31.5) | 63 (75.9) |  |
| Unemployed | 5 (3.8) | 0 (0) |  |
| Unable to work or disabled | 14 (10.8) | 0 (0) |  |
| Student | 1 (0.8) | 0 (0) |  |
| If other, please specify your employment | 3 (2.3) | 6 (7.2) |  |
| Don't know/Prefer not to answer | 0 (0) | 0 (0) |  |
| Missing | 0 (0) | 1 (1.2) |  |
| **Total Family Income** |  |  | 0.904 |
| \$60,000 or more per year | 76 (58.5) | 44 (53.0) | |
| \$30,000 to \$60,000 per year | 17 (13.0) | 13 (15.7) | |
| \$10,000 to \$30,000 per year | 10 (7.7) | 5 (6.0) | |
| Less than \$10,000 per year | 1 (0.8) | 1 (1.2) | |
| Don't know/Prefer not to answer | 26 (20.0) | 19 (22.9) |  |
| Missing | 0 (0) | 1 (1.2) |  |
| **Health care coverage** |  |  | <0.001 |
| Medicare | 19 (14.6) | 64 (77.1) |  |
| Employer/union-provided health insurance<br>(including plans purchased through another person's<br>employer) | 73 (56.2) | 12 (14.5) |  |
| A plan that you or another family member buys on<br>your own | 14 (10.7) | 1 (1.2) |  |
| TRICARE (formerly CHAMPUS), VA, or Military | 8 (6.2) | 4 (4.8) |  |
| None (no coverage) | 7 (5.4) | 0 (0) |  |
| Other (specify): | 4 (3.1) | 2 (2.4) |  |
| Medicaid or other state program | 3 (2.3) | 0 (0) |  |
| Indian Health Service | 0 (0) | 0 (0) |  |
| Don't Know/Prefer not to answer | 2 (1.5) | 0 (0) |  |

\*5 participants did not disclose their age

**Supplemental Table 3.** Demographics by race/ethnicity.

| **Supplemental Table 3. Demographics by race/ethnicity** |  |  |  |
| --- | --- | --- | --- |
| **Characteristic** | **White Non-Hispanic**<br>**(N=175)** | **Everyone else**<br>**(N=31)** | **P-value** |
| **Age, mean (SD)\*** | 64.1 (8.3) | 62.4 (7.3) | 0.304 |
| **Sex assigned at birth** |  |  | 0.650 |
| Female | 132 (75.4) | 25 (80.6) |  |
| Male | 43 (24.6) | 6 (19.4) |  |
| Don't Know or Prefer Not to Answer | 0 (0) | 0 (0) |  |
| **Current gender identity** |  |  | 0.704 |
| Female | 129 (73.7) | 25 (80.6) |  |
| Male | 41 (23.4) | 6 (19.4) |  |
| I use a different term (please specify): | 1 (0.6) | 0 (0) |  |
| Transgender | 0 (0) | 0 (0) |  |
| Non-binary | 0 (0) | 0 (0) |  |
| Two-spirit | 0 (0) | 0 (0) |  |
| Don't know or prefer not to answer | 0 (0) | 0 (0) |  |
| Missing | 4 (2.3) | 0 (0) |  |
| **Highest level of education** |  |  | 0.905 |
| No formal schooling | 0 (0) | 0 (0) |  |
| Less than 8th grade | 0 (0) | 0 (0) |  |
| Some high school | 0 (0) | 0 (0) |  |
| High school graduate/GED | 12 (6.9) | 2 (6.5) |  |
| Technical/vocational training | 13 (7.4) | 1 (3.2) |  |
| Associate degree/some college | 37 (21.1) | 8 (25.8) |  |
| Bachelor's degree | 43 (24.6) | 9 (29.0) |  |
| Post-graduate degree | 70 (40.0) | 11 (35.5) |  |
| Do Not Know or Prefer Not to Answer | 0 (0) | 0 (0) |  |
| **Have you smoked at least 100 cigarettes in your entire life?** |  |  | 0.470 |
| Yes | 107 (61.1) | 15 (48.4) |  |
| No | 39 (22.3) | 8 (25.8) |  |
| Never smoked | 26 (14.9) | 8 (25.8) |  |
| Don't know | 1 (0.6) | 0 (0) |  |
| Prefer not to answer | 2 (1.1) | 0 (0) |  |
| **Current Smoker** |  |  | 0.695 |
| No | 131 (74.9) | 22 (71.0) |  |
| Yes | 13 (7.4) | 1 (3.2) |  |
| Missing | 31 (17.7) | 8 (25.8) |  |
| **Veteran status (select all that apply) – required question** |  |  | 0.118 |
| I am NOT a veteran | 154 (88.0) | 27 (87.0) |  |
| Retired or separated veteran | 14 (8.0) | 2 (6.5) |  |
| Disabled veteran | 6 (3.4) | 0 (0) |  |
| Retired or separated veteran, and Disabled veteran | 1 (0.6) | 2 (6.5) |  |
| **Marital Status** |  |  | **0.001** |
| Married | 124 (70.9) | 13 (42.0) |  |
| Divorced | 21 (12.0) | 10 (32.3) |  |
| Single | 13 (7.4) | 5 (16.1) |  |
| Widowed | 11 (6.3) | 1 (3.2) |  |
| Living with significant other | 6 (3.4) | 0 (0) |  |

**Supplemental Table 3. Continued**
| Characteristic | White Non-Hispanic<br>(N=175) | Everyone else<br>(N=31) | P-value |
| --- | --- | --- | --- |
| Prefer not to answer | 0 (0) | 1 (3.2) |  |
| Separated | 0 (0) | 1 (3.2) |  |
| **Employment Status** |  |  | 0.167 |
| Employed | 56 (32.0) | 18 (58.1) |  |
| Retired | 94 (53.7) | 12 (38.7) |  |
| Unemployed | 4 (2.3) | 0 (0) |  |
| Unable to work or disabled | 12 (6.8) | 1 (3.2) |  |
| Student | 1 (0.6) | 0 (0) |  |
| If other, please specify your employment | 7 (4.0) | 0 (0) |  |
| Don't know/Prefer not to answer | 0 (0) | 0 (0) |  |
| Missing | 1 (0.6) | 0 (0) |  |
| **Total Family Income** |  |  | 0.294 |
| \$60,000 or more per year | 99 (56.6) | 14 (45.2) | |
| \$30,000 to \$60,000 per year | 24 (13.7) | 6 (19.3) | |
| \$10,000 to \$30,000 per year | 11 (6.3) | 5 (16.1) | |
| Less than \$10,000 per year | 2 (1.1) | 0 (0) | |
| Don't know/Prefer not to answer | 38 (21.7) | 6 (19.4) |  |
| Missing | 1 (0.6) | 0 (0) |  |
| **Health care coverage -- required question** |  |  | 0.236 |
| Medicare | 70 (40.0) | 10 (32.3) |  |
| Employer/union-provided health insurance (including plans purchased through another person's employer) | 69 (39.4) | 11 (35.4) |  |
| A plan that you or another family member buys on your own | 14 (8.0) | 1 (3.2) |  |
| TRICARE (formerly CHAMPUS), VA, or Military | 8 (4.6) | 4 (12.9) |  |
| None (no coverage) | 4 (2.3) | 2 (6.5) |  |
| Other (specify): | 5 (2.9) | 2 (6.5) |  |
| Medicaid or other state program | 3 (1.7) | 1 (3.2) |  |
| Indian Health Service | 0 (0) | 0 (0) |  |
| Don't Know/Prefer not to answer | 2 (1.1) | 0 (0) |  |

\*5 participants did not disclose their age

**Supplemental Table 4.** Demographics by veteran status.

| **Supplemental Table 4. Demographics by veteran status** |  |  |  |
| --- | --- | --- | --- |
| **Characteristic** | **Not a Veteran**<br>**(N=192)** | **Veteran**<br>**(N=26)** | **P-value** |
| **Age, mean (SD)\*** | 63.7 (7.9) | 65.5 (9.8) | 0.307 |
| **Race** |  |  | 0.888 |
| White or Caucasian | 171 (89.1) | 24 (92.3) |  |
| Black or African American | 9 (4.6) | 2 (7.7) |  |
| Asian | 3 (1.6) | 0 (0) |  |
| Hawaiian or Pacific Islander | 0 (0) | 0 (0) |  |
| American Indian or Alaska Native | 0 (0) | 0 (0) |  |
| American Indian, Black, and White | 1 (0.5) | 0 (0) |  |
| Black and Other | 1 (0.5) | 0 (0) |  |
| Don't know/Prefer not to answer | 3 (1.6) | 0 (0) |  |
| Something else/Not Sure (please specify): | 1 (0.5) | 0 (0) |  |
| Missing | 3 (1.6) | 0 (0) |  |
| **Hispanic, Latino, or Spanish origin** |  |  | 0.199 |
| No not of Hispanic, Latino or Spanish origin | 166 (86.4) | 23 (88.5) |  |
| Yes, Puerto Rican | 7 (3.6) | 0 (0) |  |
| Yes, another Hispanic, Latino or Spanish origin (please specify): | 4 (2.1) | 0 (0) |  |
| Yes, Cuban | 3 (1.6) | 0 (0) |  |
| Yes, Mexican, Mexican American, Chicano/a | 1 (0.5) | 2 (7.7) |  |
| Don't Know or Prefer Not to Answer | 3 (1.6) | 0 (0) |  |
| Missing | 8 (4.2) | 1 (3.8) |  |
| **Sex assigned at birth** |  |  | <0.001 |
| Female | 158 (82.3) | 10 (38.5) |  |
| Male | 34 (17.7) | 16 (61.5) |  |
| Don't Know or Prefer Not to Answer | 0 (0) | 0 (0) |  |
| **Current gender identity** |  |  | <0.001 |
| Female | 155 (80.7) | 10 (38.5) |  |
| Male | 33 (17.2) | 15 (57.7) |  |
| I use a different term (please specify): | 0 (0) | 1 (3.8) |  |
| Transgender | 0 (0) | 0 (0) |  |
| Non-binary | 0 (0) | 0 (0) |  |
| Two-spirit | 0 (0) | 0 (0) |  |
| Don't know or prefer not to answer | 0 (0) | 0 (0) |  |
| Missing | 4 (2.1) | 0 (0) |  |
| **Highest level of education** |  |  | 0.300 |
| No formal schooling | 0 (0) | 0 (0) |  |
| Less than 8th grade | 0 (0) | 0 (0) |  |
| Some high school | 0 (0) | 0 (0) |  |
| High school graduate/GED | 13 (6.8) | 3 (11.5) |  |
| Technical/vocational training | 13 (6.8) | 2 (7.7) |  |
| Associate degree/some college | 43 (22.4) | 4 (15.4) |  |
| Bachelor's degree | 45 (23.4) | 10 (38.4) |  |
| Post-graduate degree | 78 (40.6) | 7 (27.0) |  |
| Do Not Know or Prefer Not to Answer | 0 (0) | 0 (0) |  |
| **Have you smoked at least 100 cigarettes in your entire life?** |  |  | 0.112 |
| Yes | 115 (60.0) | 17 (65.4) |  |
| No | 45 (23.4) | 3 (11.6) |  |
| Never smoked | 30 (15.6) | 4 (15.4) |  |

**Supplemental Table 4. Continued**
| Characteristic |  | Not a Veteran<br>(N=192) | Veteran<br>(N=26) | P-value |
| --- | --- | --- | --- | --- |
|  | Don't know | 1 (0.5) | 1 (3.8) |  |
|  | Prefer not to answer | 1 (0.5) | 1 (3.8) |  |
| **Current Smoker** |  |  |  | 0.678 |
|  | No | 145 (75.5) | 18 (69.2) |  |
|  | Yes | 13 (6.8) | 2 (7.7) |  |
|  | Missing | 34 (17.7) | 6 (23.1) |  |
| **Marital Status** |  |  |  | 0.544 |
|  | Married | 125 (65.1) | 21 (80.8) |  |
|  | Divorced | 30 (15.6) | 3 (11.5) |  |
|  | Single | 18 (9.4) | 0 (0) |  |
|  | Widowed | 10 (5.3) | 2 (7.7) |  |
|  | Living with significant other | 6 (3.1) | 0 (0) |  |
|  | Prefer not to answer | 2 (1.0) | 0 (0) |  |
|  | Separated | 1 (0.5) | 0 (0) |  |
| **Employment Status** |  |  |  | 0.938 |
|  | Employed | 69 (35.9) | 10 (38.5) |  |
|  | Retired | 96 (50.0) | 13 (50.0) |  |
|  | Unemployed | 4 (2.1) | 1 (3.8) |  |
|  | Unable to work or disabled | 13 (6.8) | 1 (3.8) |  |
|  | Student | 1 (0.5) | 0 (0) |  |
|  | If other, please specify your employment | 8 (4.2) | 1 (3.8) |  |
|  | Don't know/Prefer not to answer | 0 (0) | 0 (0) |  |
|  | Missing | 1 (0.5) | 0 (0) |  |
| **Total Family Income** |  |  |  | 0.957 |
| | \$60,000 or more per year | 107 (55.7) | 16 (61.5) | |
| | \$30,000 to \$60,000 per year | 27 (14.2) | 3 (11.5) | |
| | \$10,000 to \$30,000 per year | 15 (7.8) | 1 (3.8) | |
| | Less than \$10,000 per year | 2 (1.0) | 0 (0) | |
|  | Don't know/Prefer not to answer | 40 (20.8) | 6 (23.2) |  |
|  | Missing | 1 (0.5) | 0 (0) |  |
| **Health care coverage** |  |  |  | 0.091 |
|  | Medicare | 78 (40.6) | 8 (30.8) |  |
|  | Employer/union-provided health insurance (including plans purchased through another person's employer) | 75 (39.1) | 10 (38.5) |  |
|  | A plan that you or another family member buys on your own | 14 (7.4) | 1 (3.8) |  |
|  | TRICARE (formerly CHAMPUS), VA, or Military | 7 (3.6) | 5 (19.2) |  |
|  | None (no coverage) | 7 (3.6) | 0 (0) |  |
|  | Other (specify): | 5 (2.6) | 2 (7.7) |  |
|  | Medicaid or other state program | 4 (2.1) | 0 (0) |  |
|  | Indian Health Service | 0 (0) | 0 (0) |  |
|  | Don't Know/Prefer not to answer | 2 (1.0) | 0 (0) |  |

\*5 participants did not disclose their age

**Supplemental Table 5.** Participant Responses by age.

| **Supplemental Table 5. Participant Responses by age** |  |  |  |
| --- | --- | --- | --- |
| **Survey Items** | **< 67**<br>**(N=130)** | **≥ 67**<br>**(N=83)** | **P-value** |
| **Preferred days and times for screening if a mobile unit visited your neighborhood** |  |  | 0.668 |
| M-F (9am-5pm) | 50 (38.4) | 44 (53.0) |  |
| Mornings, M-F (before 9am) | 3 (2.3) | 2 (2.4) |  |
| Evenings M-F (after 5pm) | 5 (3.8) | 4 (4.8) |  |
| Weekends (9am-5pm) | 14 (10.8) | 5 (6.0) |  |
| Weekends (before 9am) | 0 (0) | 0 (0) |  |
| Weekends (after 5pm) | 0 (0) | 1 (1.2) |  |
| Evenings M-F (after 5pm), and weekends (9am-5pm) | 11 (8.5) | 6 (7.3) |  |
| M-F (9am-5pm), and weekends (9am-5pm) | 10 (7.7) | 6 (7.3) |  |
| M-F (9am-5pm), Evenings M-F (after 5pm), Weekends (9am-5pm), and Weekends (after 5pm) | 3 (2.3) | 3 (3.6) |  |
| All of the above | 4 (3.1) | 4 (4.8) |  |
| Infrequent Responses | 30 (23.1) | 8 (9.6) |  |
| **Concerns about getting screened in a mobile screening unit (select all that apply)** |  |  | 0.540 |
| No Concerns | 95 (73.0) | 62 (74.8) |  |
| Accurate Results | 4 (3.1) | 3 (3.6) |  |
| Privacy | 5 (3.8) | 1 (1.2) |  |
| Cleanliness | 4 (3.1) | 1 (1.2) |  |
| Equipment | 1 (0.8) | 2 (2.4) |  |
| Temperature | 1 (0.8) | 1 (1.2) |  |
| Other | 1 (0.8) | 0 (0) |  |
| Equipment, Cleanliness, and Accurate Results | 3 (2.3) | 1 (1.2) |  |
| Equipment, Cleanliness, Temperature, Privacy, and Accurate Results | 1 (0.8) | 2 (2.4) |  |
| Infrequent Responses | 15 (11.5) | 10 (12.0) |  |
| **Comfort with a new healthcare professional conducting the screening (for example, not associated with your primary care)?** |  |  | 1.000 |
| Yes | 2 (1.5) | 1 (1.2) |  |
| No | 128 (98.5) | 82 (98.8) |  |
| **Negative perceptions concerning getting screening services mobile units? (e.g., perceived as not being able to afford proper treatment)?** |  |  | 0.842 |
| No | 83 (63.8) | 50 (60.2) |  |
| Do Not Know or Prefer Not to Answer | 24 (18.5) | 18 (21.7) |  |
| Yes | 22 (16.9) | 14 (16.9) |  |
| Missing | 1 (0.8) | 1 (1.2) |  |
| **Concerned about the cost or if your insurance covered the mobile lung cancer screening?** |  |  | 0.339 |
| No | 31 (23.8) | 21 (25.3) |  |
| Yes | 92 (70.8) | 54 (65.1) |  |
| Do Not Know or Prefer Not to Answer | 6 (4.6) | 8 (9.6) |  |
| Missing | 1 (0.8) | 0 (0) |  |
| **Likely to attend a mobile lung cancer screening event in the summer?** |  |  | 0.142 |
| Extremely Unlikely | 55 (42.3) | 23 (27.7) |  |
| Unlikely | 35 (26.9) | 26 (31.3) |  |
| Neutral | 27 (20.8) | 18 (21.7) |  |
| Likely | 10 (7.7) | 12 (14.5) |  |
| Extremely Likely | 1 (0.8) | 2 (2.4) |  |
| Missing | 2 (1.5) | 2 (2.4) |  |

| **Supplemental Table 5. Continued** |  |  |  |
| --- | --- | --- | --- |
| **Survey Items** | **< 67**<br>**(N=130)** | **≥ 67**<br>**(N=83)** | **P-value** |
| **Good time of year for a mobile lung cancer screening event** |  |  | 0.275 |
| No Preference | 75 (57.7) | 40 (48.2) |  |
| Spring | 6 (4.6) | 3 (3.7) |  |
| Summer | 5 (3.8) | 2 (2.4) |  |
| Fall | 10 (7.7) | 9 (10.8) |  |
| Winter | 7 (5.4) | 10 (12.0) |  |
| Winter, Spring and Fall | 11 (8.5) | 6 (7.2) |  |
| Winter and Fall | 5 (3.8) | 3 (3.7) |  |
| Winter and Spring | 2 (1.5) | 4 (4.8) |  |
| Winter, Spring, Summer, and Fall | 1 (0.8) | 5 (6.0) |  |
| Summer and Fall | 2 (1.5) | 1 (1.2) |  |
| Spring and Fall | 3 (2.3) | 0 (0) |  |
| Spring and Summer | 1 (0.8) | 0 (0) |  |
| Spring, Summer, and Fall | 1 (0.8) | 0 (0) |  |
| Winter, Summer, and Fall | 1 (0.8) | 0 (0) |  |
| **Length of time willing to wait for a screening** |  |  | 0.993 |
| Less than 15 | 30 (23.0) | 18 (21.7) |  |
| 15-30 minutes | 76 (58.5) | 50 (60.2) |  |
| 30-60 minutes | 21 (16.2) | 13 (15.7) |  |
| More than 60 minutes | 3 (2.3) | 2 (2.4) |  |
| **Where would you like the waiting room/area to be located?** |  |  | 0.462 |
| Indoor | 84 (64.6) | 45 (54.2) |  |
| Outdoor | 6 (4.6) | 4 (4.8) |  |
| No Preference | 26 (20.0) | 21 (25.3) |  |
| Other | 13 (10.0) | 12 (14.5) |  |
| Missing | 1 (0.8) | 1 (1.2) |  |
| **\*\*Easiest location for mobile bus screening** |  |  | 0.478 |
| No preference | 44 (33.8) | 30 (36.2) |  |
| Neighborhood | 8 (6.2) | 8 (9.6) |  |
| Shopping mall/grocery store | 6 (4.7) | 3 (3.6) |  |
| Downtown in my city | 2 (1.5) | 2 (2.4) |  |
| Fire station or police station | 2 (1.5) | 0 (0) |  |
| Church and other religious institutions | 0 (0) | 1 (1.2) |  |
| School | 1 (0.8) | 0 (0) |  |
| Health fair | 0 (0) | 0 (0) |  |
| Post office | 0 (0) | 0 (0) |  |
| Neighborhood, and shopping mall/grocery store | 6 (4.7) | 3 (3.6) |  |
| Neighborhood, shopping mall/grocery store, and Church or other religious institutions | 5 (3.8) | 1 (1.2) |  |
| Neighborhood, Health Fair, and Church or other religious institutions | 2 (1.5) | 2 (2.4) |  |
| Other | 2 (1.5) | 0 (0) |  |
| Infrequent Responses | 52 (40.0) | 33 (39.8) |  |
| **Has your healthcare provider/doctor ever talked to you about getting screened for lung cancer?** |  |  | 0.811 |
| No | 77 (59.2) | 47 (56.6) |  |
| Yes | 40 (30.8) | 29 (35.0) |  |
| Don't recall/ Don't remember | 13 (10.0) | 7 (8.4) |  |
| **Survey Items** | **< 67**<br>**(N=130)** | **≥ 67**<br>**(N=83)** | **P-value** |
| **\*\*Previous experiences with cancer screenings?** |  |  | **0.387** |
| Breast | 12 (9.2) | 3 (3.6) |  |
| Prostate | 2 (1.5) | 3 (3.6) |  |
| Colon | 2 (1.5) | 2 (2.4) |  |
| Lung | 3 (2.4) | 1 (1.2) |  |
| Skin | 0 (0) | 2 (2.4) |  |
| Cervical | 0 (0) | 0 (0) |  |
| Other | 2 (1.5) | 1 (1.2) |  |
| No previous experiences with cancer screenings | 2 (1.5) | 1 (1.2) |  |
| Breast, Colon, Skin, and Cervical | 24 (18.5) | 11 (13.4) |  |
| Breast, Colon, Lung, Skin, and Cervical | 9 (6.9) | 7 (8.4) |  |
| Breast, Colon, and Skin | 9 (6.9) | 3 (3.6) |  |
| Colon, Prostate, and Skin | 3 (2.4) | 8 (9.6) |  |
| Breast, Colon, and Cervical | 6 (4.6) | 2 (2.4) |  |
| Breast, Skin, and Cervical | 7 (5.4) | 1 (1.2) |  |
| Infrequent Responses | 49 (37.7) | 38 (45.8) |  |
| **Do you trust the health care system (for example, hospitals, pharmacies, and other organizations involved in health care)?** |  |  | **0.303** |
| Very | 67 (51.5) | 49 (59.0) |  |
| Somewhat | 54 (41.5) | 32 (38.6) |  |
| A little | 9 (7.0) | 2 (2.4) |  |
| Not at all | 0 (0) | 0 (0) |  |
| **Likely to get screened for lung cancer if you could get a voucher or insurance to pay for it?** |  |  | **0.003** |
| Extremely likely | 91 (70.0) | 41 (49.4) |  |
| Likely | 25 (19.2) | 22 (26.5) |  |
| Neutral | 11 (8.5) | 16 (19.3) |  |
| Extremely Unlikely | 0 (0) | 3 (3.6) |  |
| Unlikely | 1 (0.8) | 1 (1.2) |  |
| Missing | 2 (1.5) | 0 (0) |  |
| **Would you be more likely to get screened for lung cancer if the screening could be done close to your home or in your neighborhood?** |  |  | **0.131** |
| Very likely | 81 (62.3) | 42 (50.6) |  |
| Likely | 33 (25.4) | 28 (33.7) |  |
| Neutral | 16 (12.3) | 11 (13.3) |  |
| Extremely Unlikely | 0 (0) | 2 (2.4) |  |
| Unlikely | 0 (0) | 0 (0) |  |
| **Would you be more likely to be screened for lung cancer if your doctor/health care provider recommended it?** |  |  | **0.358** |
| Very Likely | 83 (63.8) | 57 (68.7) |  |
| Likely | 30 (23.1) | 17 (20.5) |  |
| Neutral | 16 (12.3) | 7 (8.4) |  |
| Extremely Unlikely | 0 (0) | 2 (2.4) |  |
| Unlikely | 1 (0.8) | 0 (0) |  |
| **\*\*If you wanted more information about mobile lung cancer screening, how would you want to receive this information?** |  |  | **0.644** |
| Receiving a notification in the mail | 16 (12.3) | 10 (12.0) |  |
| Talking to your primary doctor | 11 (8.4) | 9 (10.8) |  |
| Receiving a message through your electronic health record | 7 (5.4) | 5 (6.0) |  |

**Supplemental Table 5. Continued**
| Survey Items | < 67<br>(N=130) | ≥ 67<br>(N=83) | P-value |
| --- | --- | --- | --- |
| Receiving a phone call | 3 (2.3) | 1 (1.2) |  |
| Through social media (for example, Facebook, Twitter, Instagram) | 1 (0.8) | 1 (1.2) |  |
| Reading about this in a magazine/newspaper you subscribe to | 1 (0.8) | 0 (0) |  |
| Watching a local news segment or television special about this | 1 (0.8) | 0 (0) |  |
| Other | 1 (0.8) | 3 (3.6) |  |
| Talking to your primary doctor, receiving a message through your electronic health record, and receiving a notification in the mail | 11 (8.4) | 4 (4.8) |  |
| Talking to your primary doctor, and receiving a message through your electronic health record | 6 (4.6) | 3 (3.6) |  |
| Receiving a message through your electronic health record, and receiving a notification in the mail | 5 (3.8) | 3 (3.6) |  |
| Talking to your primary doctor, and receiving notification in the mail | 4 (3.1) | 3 (3.6) |  |
| Talking to your primary doctor, receiving a phone call, receiving a message through your electronic health record, and receiving a notification in the mail | 5 (3.8) | 2 (2.4) |  |
| Receiving a phone call, receiving a message through your electronic health record, and receiving a notification in the mail | 1 (0.8) | 5 (6.0) |  |
| Infrequent Responses | 56 (43.1) | 33 (40.0) |  |
| Missing | 1 (0.8) | 1 (1.2) |  |
| **Furthest distance you would travel to get screened (mobile unit or building-hospital/clinic)?** |  |  | **0.632** |
| Less than 5 minutes | 0 (0) | 0 (0) |  |
| 5-10 minutes | 13 (10.0) | 11 (13.3) |  |
| 10-20 minutes | 62 (47.7) | 35 (42.2) |  |
| More than 20 minutes | 55 (42.3) | 37 (44.5) |  |
| **Would you be comfortable talking with your doctor about lung cancer screening in a virtual appointment?** |  |  | **0.988** |
| Very comfortable | 89 (68.5) | 57 (68.7) |  |
| Somewhat comfortable | 17 (13.1) | 12 (14.5) |  |
| Neutral | 16 (12.3) | 9 (10.8) |  |
| Somewhat uncomfortable | 1 (0.8) | 0 (0) |  |
| Very uncomfortable | 7 (5.3) | 4 (4.8) |  |
| Missing | 0 (0) | 1 (1.2) |  |
| **If you needed a follow-up appointment after your lung cancer screening, where would you be willing to have it?** |  |  | **0.011** |
| Hospital/clinic | 20 (15.3) | 28 (33.8) |  |
| Mobile screening unit | 0 (0) | 0 (0) |  |
| Virtual | 1 (0.8) | 1 (1.2) |  |
| Other | 4 (3.1) | 5 (6.0) |  |
| Hospital/Clinic, and Mobile Screening Unit | 7 (5.4) | 5 (6.0) |  |
| Hospital/Clinic, Mobile Screening Unit, and Virtual | 91 (70.0) | 38 (45.8) |  |
| Hospital/Clinic and Virtual | 5 (3.8) | 4 (4.8) |  |
| Mobile Screening Unit, and Other | 1 (0.8) | 0 (0) |  |
| Virtual, and Other | 1 (0.8) | 0 (0) |  |
| Hospital/clinic, Virtual, and Other | 0 (0) | 0 (0) |  |
| All the above, and Other | 0 (0) | 1 (1.2) |  |
| Hospital/Clinic, Mobile Screening Unit, and Other | 0(0) | 1 (1.2) |  |

\*Fisher's exact test was used to compare age, except where denoted with \*\*

**Supplemental Table 6.** Participant Responses by race/ethnicity.

| **Supplemental Table 6. Participant Responses by race/ethnicity** |  |  |  |
| --- | --- | --- | --- |
| **Survey Items** | **White Non-Hispanic**<br>**(N=175)** | **Everyone else**<br>**(N=31)** | **P-value** |
| **Preferred days and times for screening if a mobile unit visited your neighborhood** |  |  | 0.187 |
| M-F (9am-5pm) | 79 (45.1) | 13 (41.9) |  |
| Mornings, M-F (before 9am) | 4 (2.3) | 1 (3.2) |  |
| Evenings M-F (after 5pm) | 9 (5.1) | 0 (0) |  |
| Weekends (9am-5pm) | 12 (6.9) | 7 (22.6) |  |
| Weekends (before 9am) | 0 (0) | 0 (0) |  |
| Weekends (after 5pm) | 1 (0.6) | 0 (0) |  |
| Evenings M-F (after 5pm), and weekends (9am-5pm) | 14 (8.0) | 1 (3.2) |  |
| M-F (9am-5pm), and weekends (9am-5pm) | 13 (7.4) | 2 (6.5) |  |
| M-F (9am-5pm), Evenings M-F (after 5pm), Weekends (9am-5pm), and Weekends (after 5pm) | 7 (4.0) | 0 (0) |  |
| All of the above | 7 (4.0) | 0 (0) |  |
| Infrequent Responses | 29 (16.6) | 7 (22.6) |  |
| **Concerns about getting screened in a mobile screening unit** |  |  | 0.114 |
| No Concerns | 132 (75.4) | 19 (61.2) |  |
| Accurate Results | 7 (4.0) | 1 (3.2) |  |
| Privacy | 5 (2.9) | 1 (3.2) |  |
| Cleanliness | 2 (1.1) | 3 (9.7) |  |
| Equipment | 1 (0.6) | 2 (6.5) |  |
| Temperature | 2 (1.1) | 0 (0) |  |
| Other | 1 (0.6) | 0 (0) |  |
| Equipment, Cleanliness, and Accurate Results | 4 (2.3) | 0 (0) |  |
| Equipment, Cleanliness, Temperature, Privacy, and Accurate Results | 2 (1.1) | 2 (6.5) |  |
| Infrequent Responses | 19 (10.9) | 3 (9.7) |  |
| **Comfort with a new healthcare professional conducting the screening (for example, not associated with your primary care)?** |  |  | 1.000 |
| Yes | 173 (98.9) | 31 (100.0) |  |
| No | 2 (1.1) | 0 (0) |  |
| **Negative perceptions concerning getting screening services mobile units? (e.g., perceived as not being able to afford proper treatment)?** |  |  | 0.959 |
| No | 109 (62.3) | 21 (67.7) |  |
| Do Not Know or Prefer Not to Answer | 35 (20.0) | 6 (19.4) |  |
| Yes | 29 (16.6) | 4 (12.9) |  |
| Missing | 2 (1.1) | 0 (0) |  |
| **Concerned about the cost or if your insurance covered the mobile lung cancer screening?** |  |  | 0.018 |
| No | 49 (28.0) | 2 (6.5) |  |
| Yes | 114 (65.1) | 26 (83.9) |  |
| Do Not Know or Prefer Not to Answer | 11 (6.3) | 3 (9.7) |  |
| Missing | 1 (0.6) | 0 (0) |  |
| **Likely to attend a mobile lung cancer screening event in the summer?** |  |  | 0.851 |
| Extremely Unlikely | 64 (36.6) | 13 (41.9) |  |
| Unlikely | 47 (26.9) | 10 (32.3) |  |
| Neutral | 39 (22.3) | 5 (16.1) |  |
| Likely | 19 (10.8) | 2 (6.5) |  |
| Extremely Likely | 3 (1.7) | 0 (0) |  |
| Missing | 3 (1.7) | 1 (3.2) |  |

**Supplemental Table 6. Continued**
| Survey Items | White Non-Hispanic<br>(N=175) | Everyone else<br>(N=31) | P-value |
| --- | --- | --- | --- |
| **Good time of year for a mobile lung cancer screening event** |  |  | **0.005** |
| No Preference | 103 (58.9) | 11 (35.5) |  |
| Spring | 7 (4.0) | 2 (6.5) |  |
| Summer | 4 (2.3) | 4 (12.9) |  |
| Fall | 12 (6.9) | 6 (19.4) |  |
| Winter | 15 (8.6) | 0 (0) |  |
| Winter, Spring and Fall | 13 (7.4) | 3 (9.7) |  |
| Winter and Fall | 5 (2.9) | 1 (3.2) |  |
| Winter and Spring | 5 (2.9) | 1 (3.2) |  |
| Winter, Spring, Summer, and Fall | 6 (3.4) | 0 (0) |  |
| Summer and Fall | 2 (1.1) | 1 (3.2) |  |
| Spring and Fall | 2 (1.1) | 1 (3.2) |  |
| Spring and Summer | 0 (0) | 0 (0) |  |
| Spring, Summer, and Fall | 0 (0) | 1 (3.2) |  |
| Winter, Summer, and Fall | 1 (0.5) | 0 (0) |  |
| **Length of time willing to wait for a screening** |  |  | **0.858** |
| Less than 15 | 39 (22.3) | 7 (22.6) |  |
| 15-30 minutes | 105 (60.0) | 18 (58.1) |  |
| 30-60 minutes | 28 (16.0) | 5 (16.1) |  |
| More than 60 minutes | 3 (1.7) | 1 (3.2) |  |
| **Where would you like the waiting room/area to be located?** |  |  | **0.389** |
| Indoor | 104 (59.4) | 23 (74.2) |  |
| Outdoor | 7 (4.0) | 0 (0) |  |
| No Preference | 41 (23.4) | 4 (12.9) |  |
| Other | 21 (12.0) | 4 (12.9) |  |
| Missing | 2 (1.2) | 0 (0) |  |
| **\*\*Easiest location for mobile bus screening** |  |  | **0.523** |
| No preference | 65 (37.1) | 8 (25.8) |  |
| Neighborhood | 12 (6.9) | 3 (9.7) |  |
| Shopping mall/grocery store | 8 (4.6) | 2 (6.5) |  |
| Downtown in my city | 3 (1.7) | 1 (3.2) |  |
| Fire station or police station | 1 (0.6) | 0 (0) |  |
| Church and other religious institutions | 1 (0.6) | 0 (0) |  |
| School | 0 (0) | 1 (3.2) |  |
| Health fair | 0 (0) | 0 (0) |  |
| Post office | 0 (0) | 0 (0) |  |
| Neighborhood, and shopping mall/grocery store | 8 (4.6) | 1 (3.2) |  |
| Neighborhood, shopping mall/grocery store, and Church or other religious institutions | 4 (2.3) | 1 (3.2) |  |
| Neighborhood, Health Fair, and Church or other religious institutions | 2 (1.1) | 2 (6.5) |  |
| Other | 2 (1.1) | 0 (0) |  |
| Infrequent Responses | 69 (39.4) | 12 (38.7) |  |
| **Has your healthcare provider/doctor ever talked to you about getting screened for lung cancer?** |  |  | **0.664** |
| No | 99 (56.6) | 20 (64.5) |  |
| Yes | 61 (34.8) | 8 (25.8) |  |
| Don't recall/ Don't remember | 15 (8.6) | 3 (9.7) |  |

| **Supplemental Table 6. Continued** |  |  |  |
| --- | --- | --- | --- |
| **Survey Items** | **White Non-Hispanic**<br>**(N=175)** | **Everyone else**<br>**(N=31)** | **P-value** |
| **\*\*Previous experiences with cancer screenings?** |  |  | **0.003** |
| Breast | 9 (5.1) | 4 (12.9) |  |
| Prostate | 3 (1.7) | 2 (6.5) |  |
| Colon | 4 (2.3) | 0 (0) |  |
| Lung | 3 (1.7) | 1 (3.2) |  |
| Skin | 2 (1.2) | 0 (0) |  |
| Cervical | 0 (0) | 0 (0) |  |
| Other | 3 (1.7) | 0 (0) |  |
| No previous experiences with cancer screenings | 2 (1.2) | 1 (3.2) |  |
| Breast, Colon, Skin, and Cervical | 33 (18.9) | 2 (6.5) |  |
| Breast, Colon, Lung, Skin, and Cervical | 16 (9.1) | 0 (0) |  |
| Breast, Colon, and Skin | 10 (5.7) | 1 (3.2) |  |
| Colon, Prostate, and Skin | 11 (6.3) | 0 (0) |  |
| Breast, Colon, and Cervical | 6 (3.4) | 3 (9.7) |  |
| Breast, Skin, and Cervical | 8 (4.6) | 0 (0) |  |
| Infrequent Responses | 65 (37.1) | 17 (54.8) |  |
| **Do you trust the health care system (for example, hospitals, pharmacies, and other organizations involved in health care)?** |  |  | **0.167** |
| Very | 101 (57.7) | 13 (42.0) |  |
| Somewhat | 64 (36.6) | 17 (54.8) |  |
| A little | 10 (5.7) | 1 (3.2) |  |
| Not at all | 0 (0) | 0 (0) |  |
| **Likely to get screened for lung cancer if you could get a voucher or insurance to pay for it?** |  |  | **0.803** |
| Extremely likely | 109 (62.4) | 20 (64.5) |  |
| Likely | 38 (21.7) | 7 (22.5) |  |
| Neutral | 24 (13.7) | 2 (6.5) |  |
| Unlikely | 2 (1.1) | 0 (0) |  |
| Extremely Unlikely | 2 (1.1) | 0 (0) |  |
| Missing | 0 (0) | 2 (6.5) |  |
| **Would you be more likely to get screened for lung cancer if the screening could be done close to your home or in your neighborhood?** |  |  | **0.782** |
| Very likely | 102 (58.3) | 16 (51.6) |  |
| Likely | 49 (28.0) | 10 (32.3) |  |
| Neutral | 22 (12.6) | 5 (16.1) |  |
| Unlikely | 0 (0) | 0 (0) |  |
| Extremely Unlikely | 2 (1.1) | 0 (0) |  |
| **Would you be more likely to be screened for lung cancer if your doctor/health care provider recommended it?** |  |  | **0.923** |
| Very Likely | 114 (65.2) | 20 (64.5) |  |
| Likely | 38 (21.7) | 8 (25.8) |  |
| Neutral | 20 (11.4) | 3 (9.7) |  |
| Unlikely | 1 (0.6) | 0 (0) |  |
| Extremely Unlikely | 2 (1.1) | 0 (0) |  |
| **\*\*If you wanted more information about mobile lung cancer screening, how would you want to receive this information?** |  |  | **0.543** |
| Receiving a notification in the mail | 20 (11.4) | 6 (19.4) |  |
| Talking to your primary doctor | 15 (8.6) | 4 (12.9) |  |

**Supplemental Table 6. Continued**
| Survey Items | White Non-Hispanic<br>(N=175) | Everyone else<br>(N=31) | P-value |
| --- | --- | --- | --- |
| Receiving a message through your electronic health record | 10 (5.7) | 3 (9.7) |  |
| Receiving a phone call | 4 (2.3) | 0 (0) |  |
| Through social media (for example, Facebook, Twitter, Instagram) | 2 (1.1) | 0 (0) |  |
| Reading about this in a magazine/newspaper you subscribe to | 0 (0) | 1 (3.2) |  |
| Watching a local news segment or television special about this | 0 (0) | 0 (0) |  |
| Other | 4 (2.3) | 0 (0) |  |
| Talking to your primary doctor, receiving a message through your electronic health record, and receiving a notification in the mail | 11 (6.3) | 2 (6.5) |  |
| Talking to your primary doctor, and receiving a message through your electronic health record | 7 (4.0) | 1 (3.2) |  |
| Receiving a message through your electronic health record, and receiving a notification in the mail | 5 (2.9) | 2 (6.5) |  |
| Talking to your primary doctor, and receiving notification in the mail | 6 (3.4) | 0 (0) |  |
| Talking to your primary doctor, receiving a phone call, receiving a message through your electronic health record, and receiving a notification in the mail | 6 (3.4) | 1 (3.2) |  |
| Receiving a phone call, receiving a message through your electronic health record, and receiving a notification in the mail | 6 (3.4) | 0 (0) |  |
| Infrequent Responses | 77 (44.1) | 11 (35.4) |  |
| Missing | 2 (1.1) | 0 (0) |  |
| **Furthest distance you would travel to get screened (mobile unit or building-hospital/clinic)?** |  |  | 0.301 |
| Less than 5 minutes | 0 (0) | 0 (0) |  |
| 5-10 minutes | 22 (12.6) | 1 (3.2) |  |
| 10-20 minutes | 74 (42.3) | 16 (51.6) |  |
| More than 20 minutes | 79 (45.1) | 14 (45.2) |  |
| **Would you be comfortable talking with your doctor about lung cancer screening in a virtual appointment?** |  |  | 0.312 |
| Very comfortable | 123 (70.2) | 19 (61.3) |  |
| Somewhat comfortable | 22 (12.6) | 6 (19.3) |  |
| Neutral | 22 (12.6) | 3 (9.7) |  |
| Somewhat uncomfortable | 0 (0) | 0 (0) |  |
| Very uncomfortable | 7 (4.0) | 3 (9.7) |  |
| Missing | 1 (0.6) | 0 (0) |  |
| **If you needed a follow-up appointment after your lung cancer screening, where would you be willing to have it?** |  |  | 0.547 |
| Hospital/Clinic, Mobile Screening Unit, and Virtual | 107 (61.1) | 17 (54.8) |  |
| Hospital/clinic | 38 (21.7) | 10 (32.3) |  |
| Mobile screening unit | 0 (0) | 0 (0) |  |
| Virtual | 2 (1.1) | 0 (0) |  |
| Other | 7 (4.0) | 1 (3.2) |  |
| Hospital/Clinic, and Mobile Screening Unit | 7 (4.0) | 3 (9.7) |  |
| Hospital/Clinic and Virtual | 9 (5.1) | 0 (0) |  |
| Mobile Screening Unit, and Other | 1 (0.6) | 0 (0) |  |
| Virtual, and Other | 1 (0.6) | 0 (0) |  |
| Hospital/clinic, Virtual, and Other | 1 (0.6) | 0 (0) |  |
| All the above, and Other | 1 (0.6) | 0 (0) |  |
| Hospital/Clinic, Mobile Screening Unit, and Other | 1 (0.6) | 0 (0) |  |

\*Fisher's exact test was used to compare race, except where denoted with \*\*

**Supplemental Table 7.**
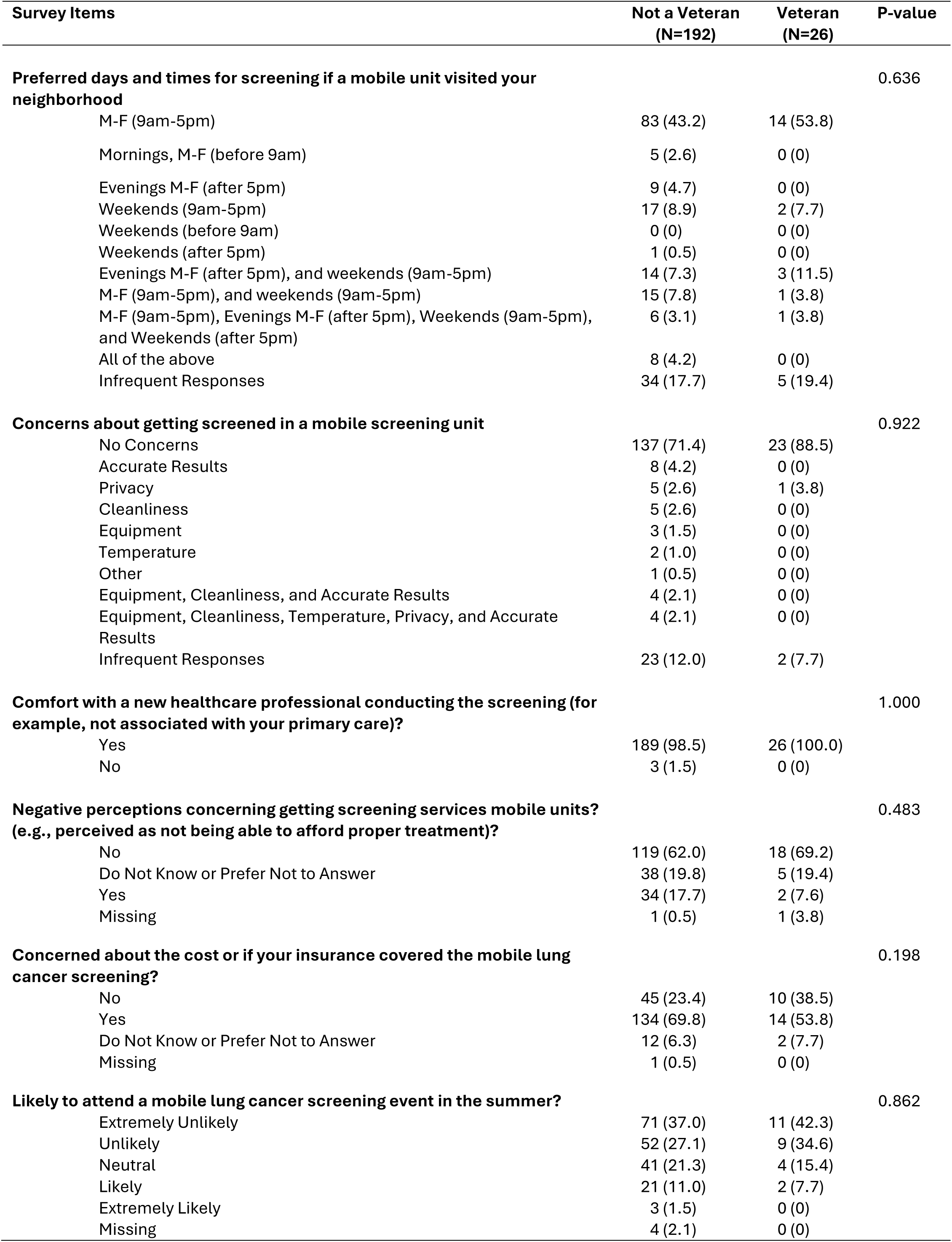
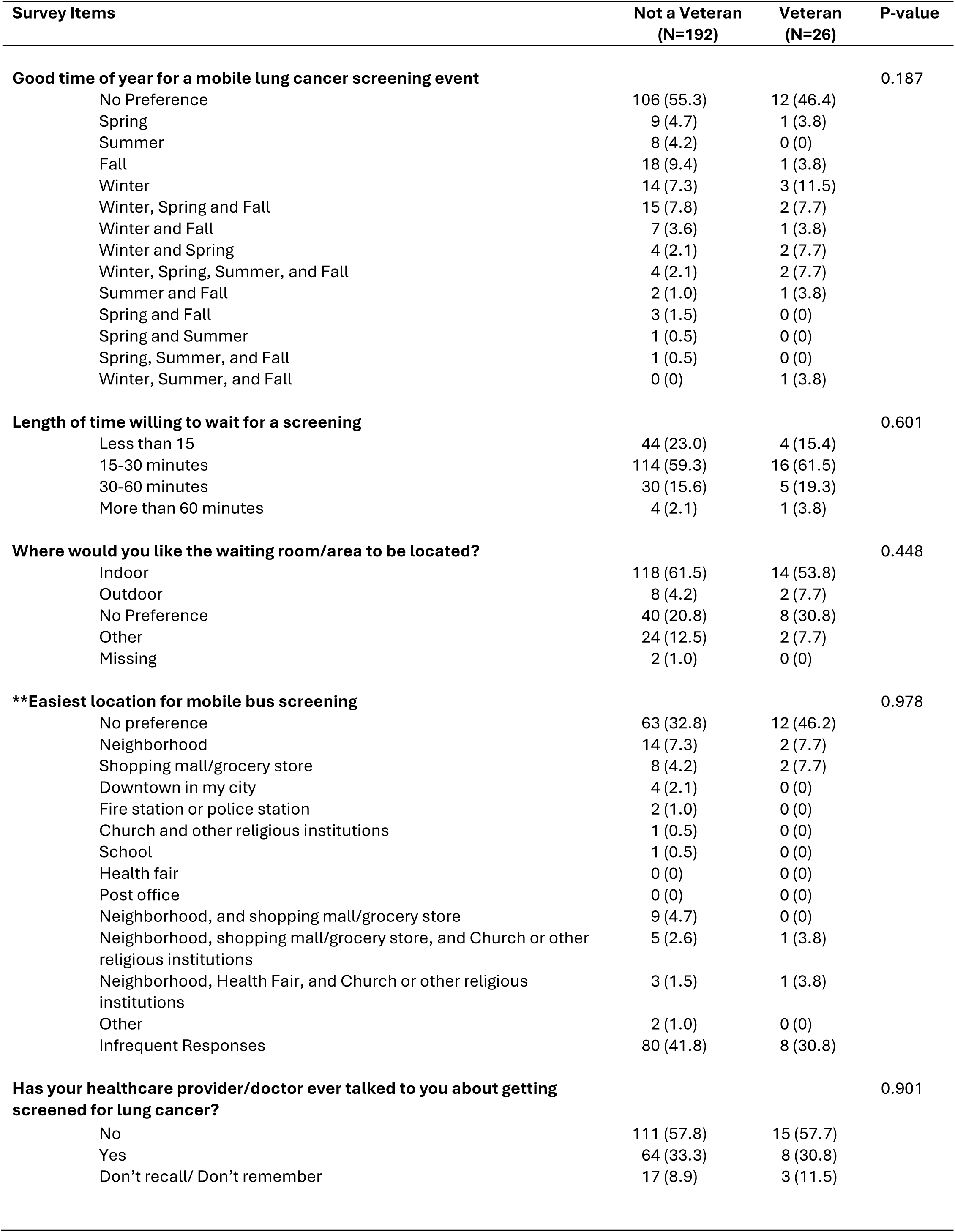
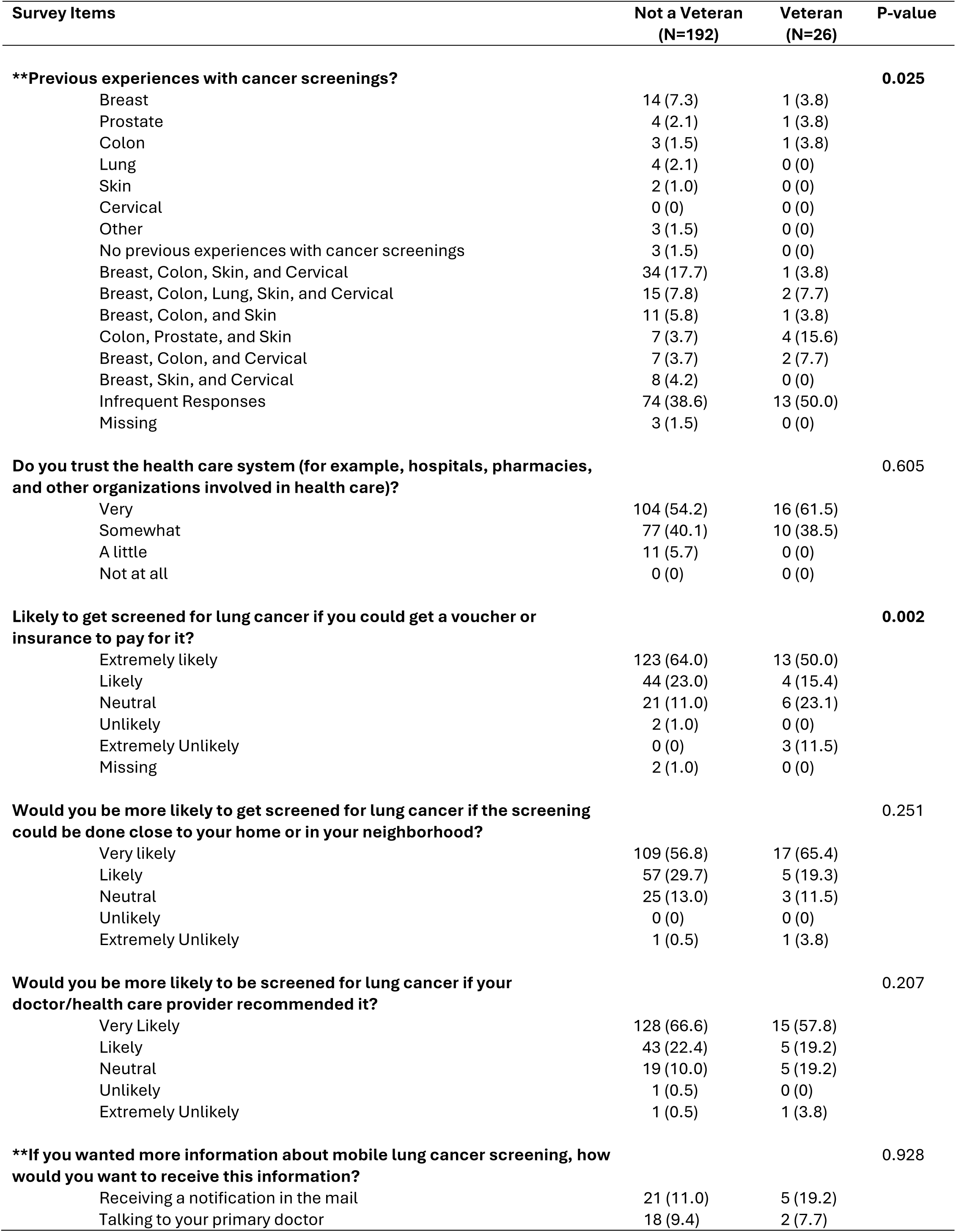
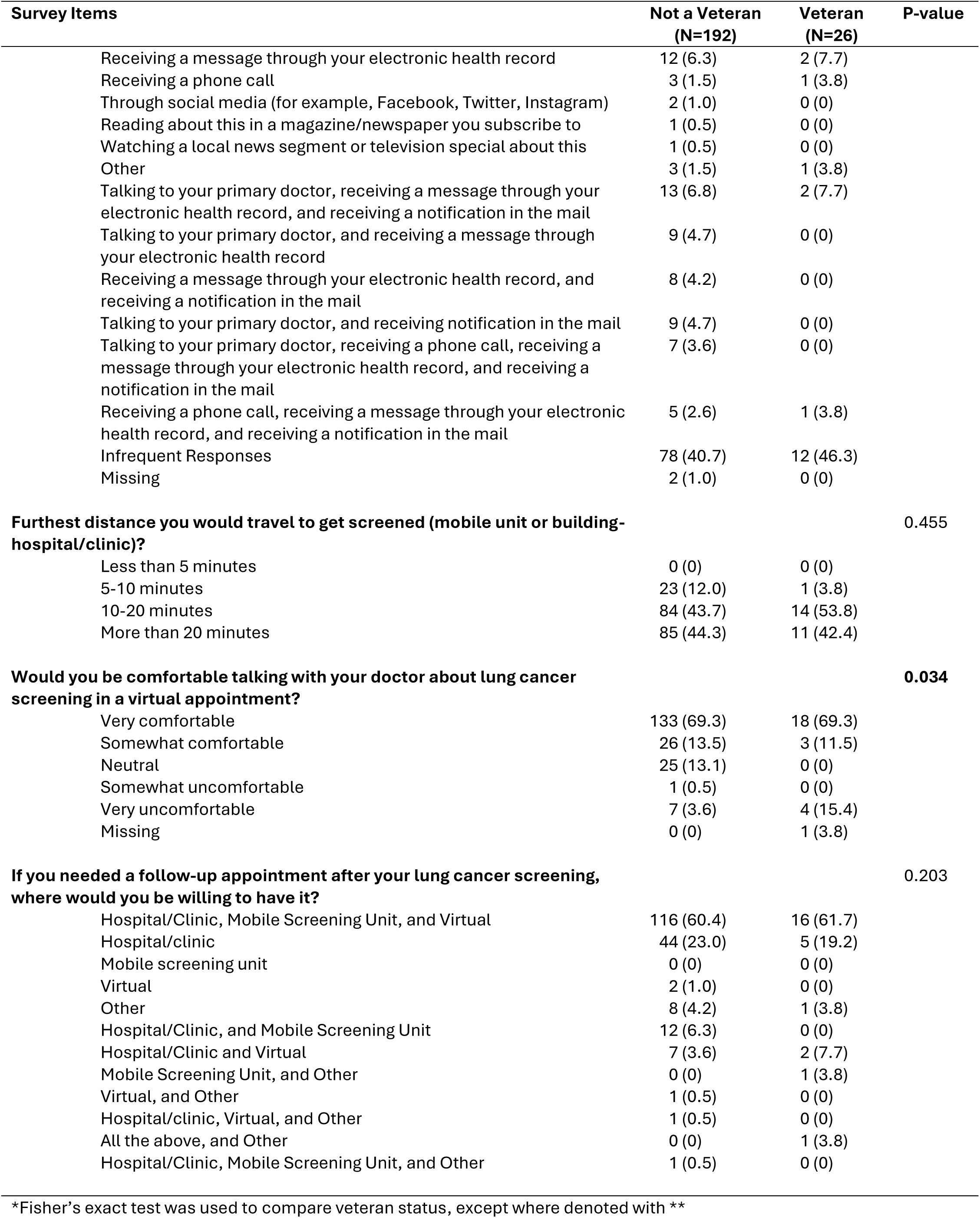
Participant Responses by veteran status.

**Supplemental Table 8.**
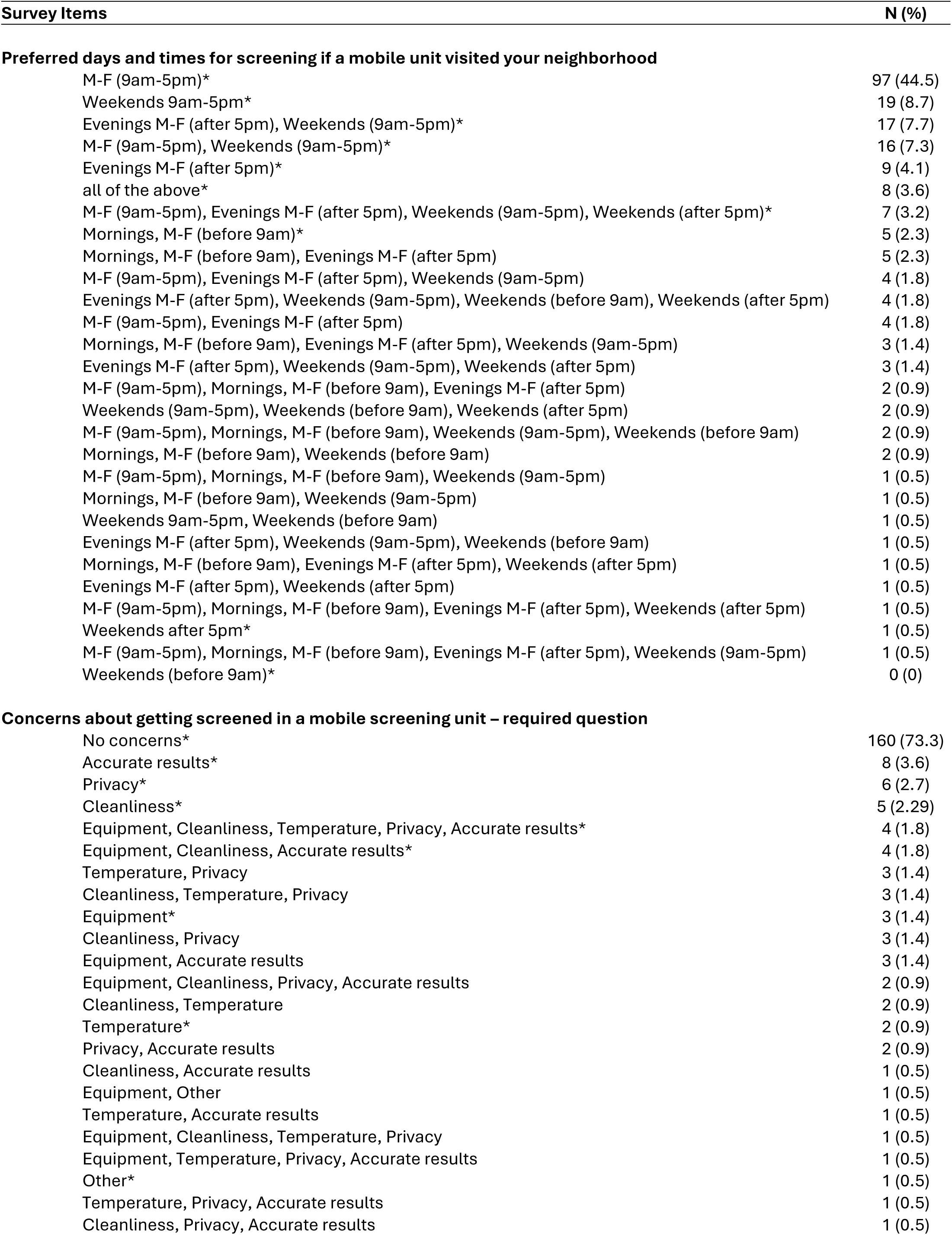
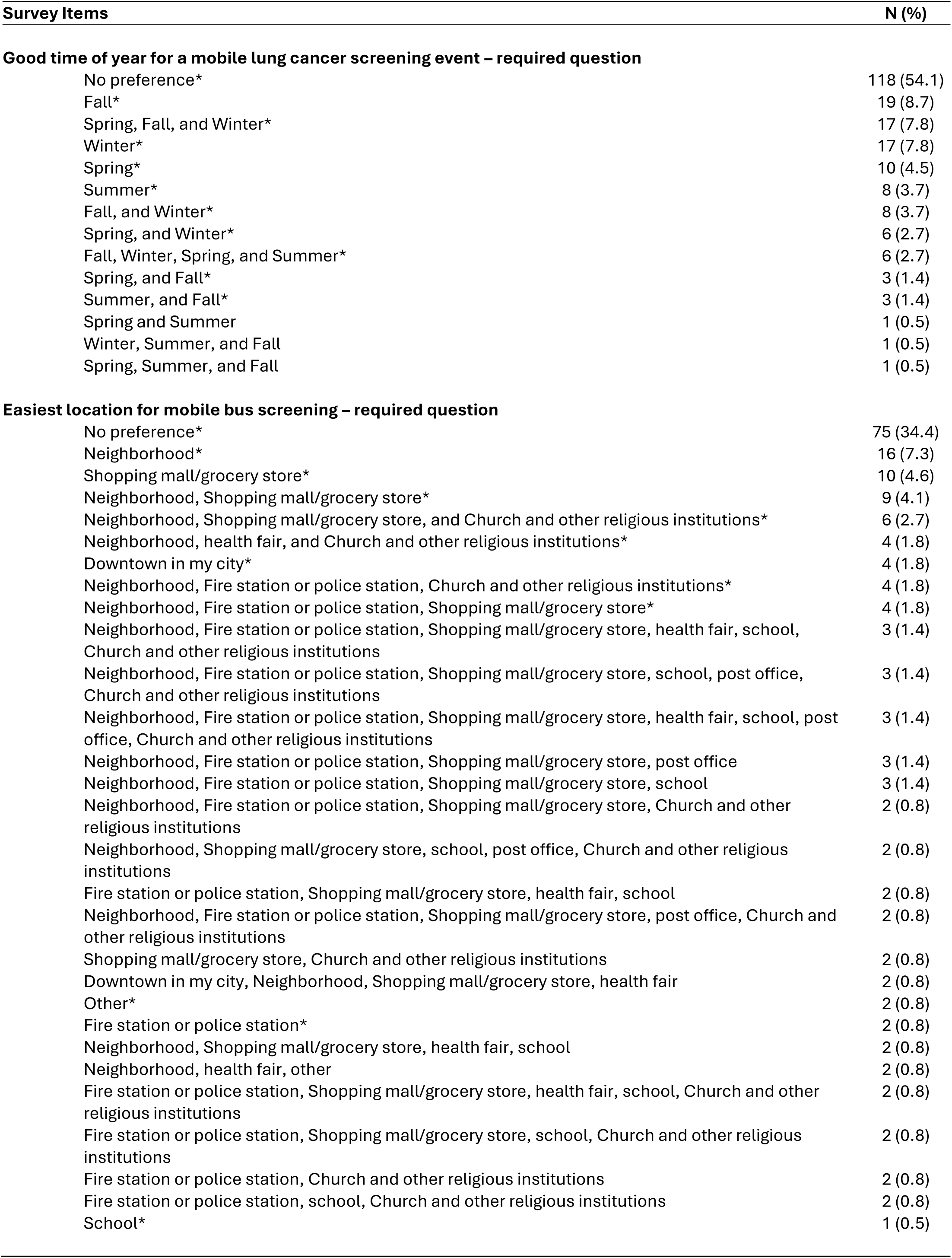
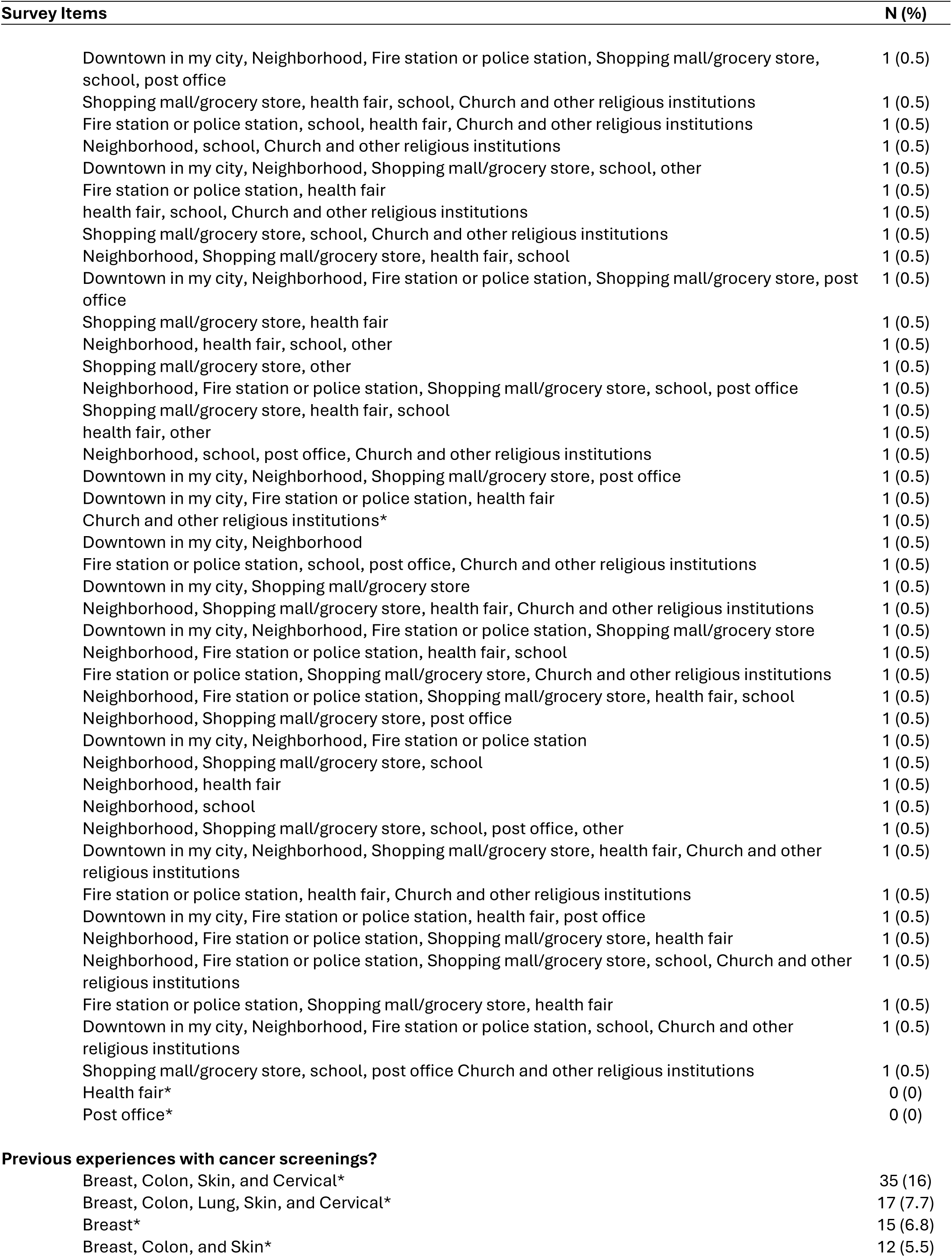
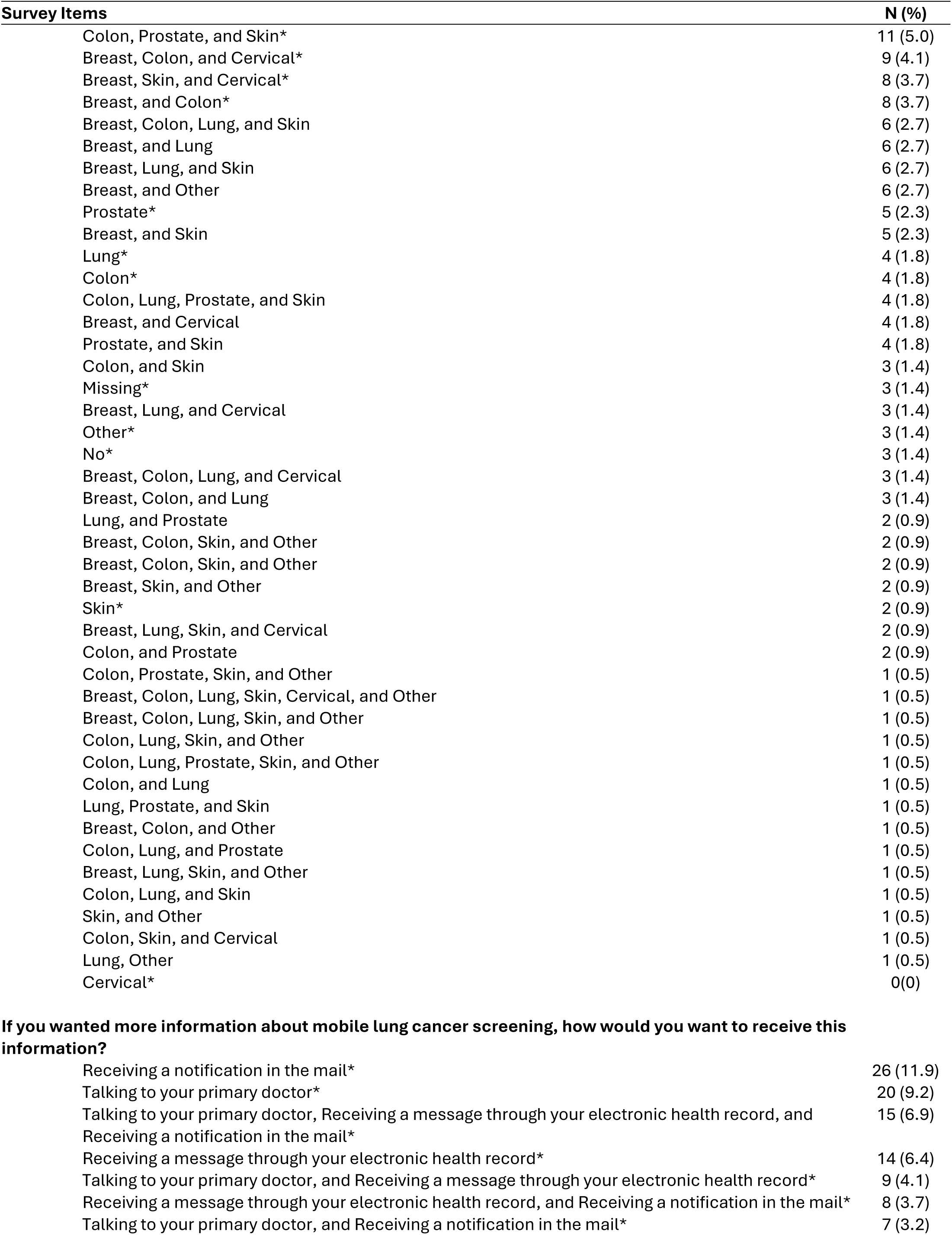
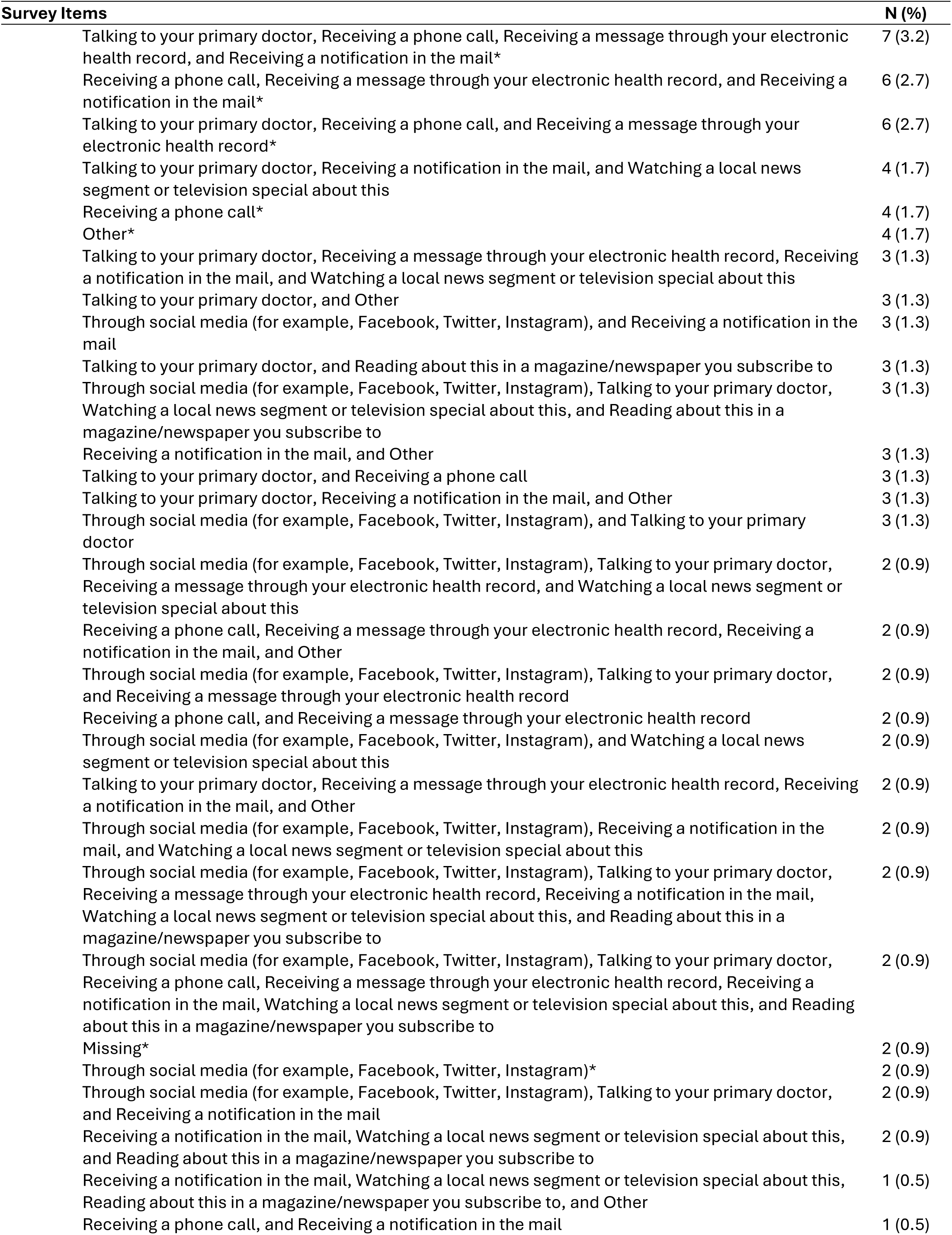
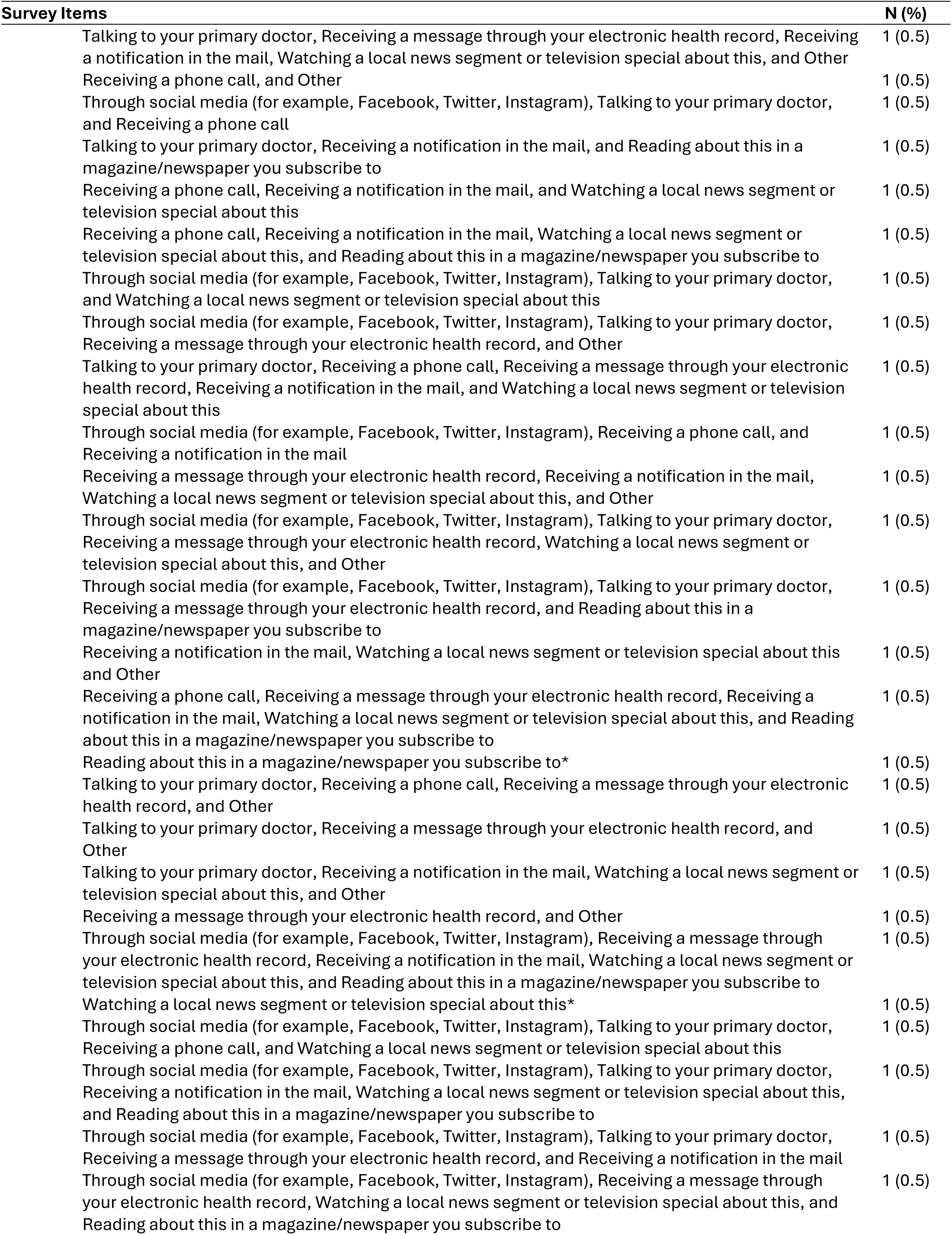
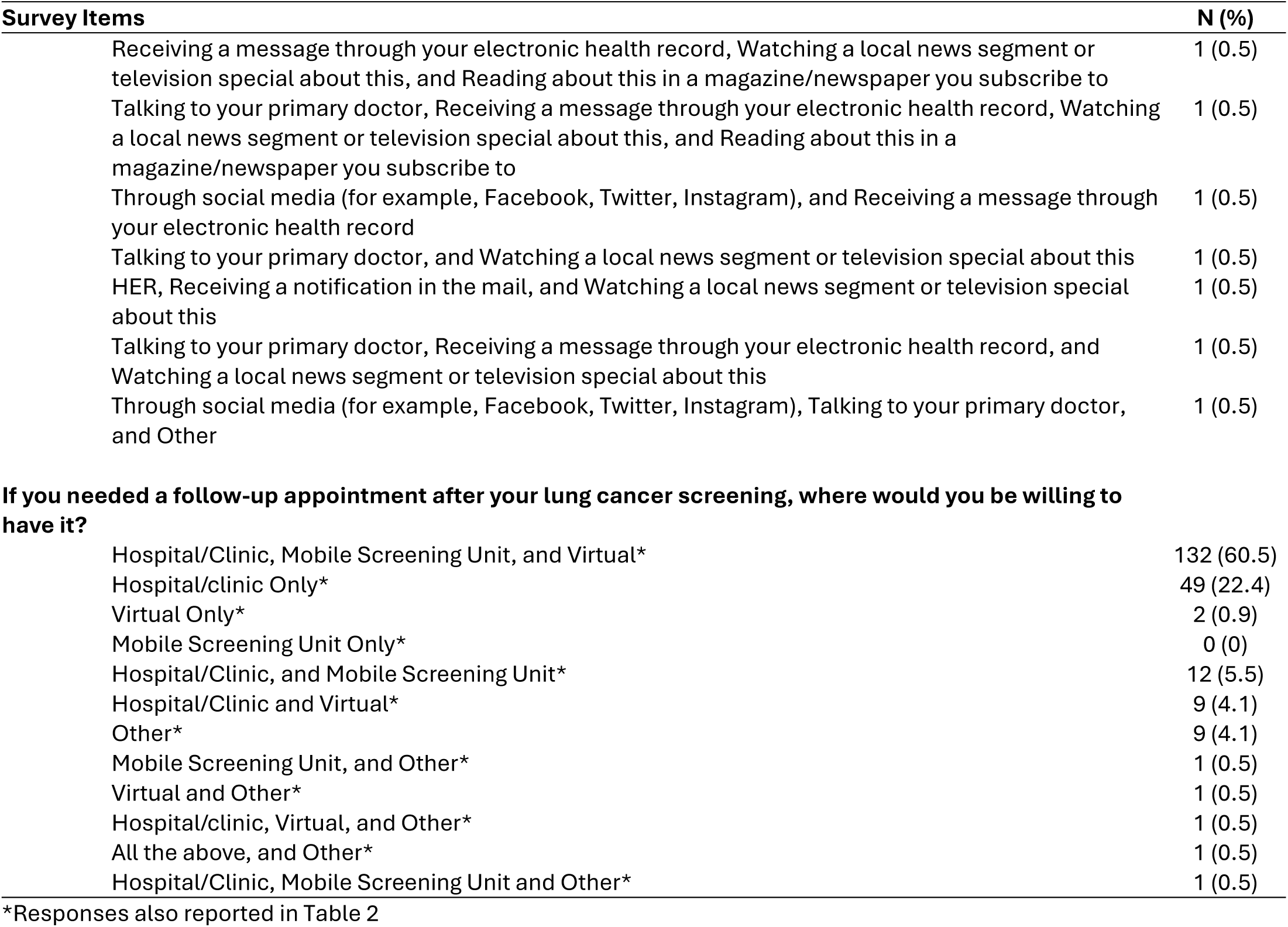
All options for “select all that apply” questions including infrequent responses.

| **Survey Items** | **N (%)** |
| --- | --- |
| **Preferred days and times for screening if a mobile unit visited your neighborhood** |  |
| M-F (9am-5pm)* | 97 (44.5) |
| Weekends 9am-5pm* | 19 (8.7) |
| Evenings M-F (after 5pm), Weekends (9am-5pm)* | 17 (7.7) |
| M-F (9am-5pm), Weekends (9am-5pm)* | 16 (7.3) |
| Evenings M-F (after 5pm)* | 9 (4.1) |
| all of the above* | 8 (3.6) |
| M-F (9am-5pm), Evenings M-F (after 5pm), Weekends (9am-5pm), Weekends (after 5pm)* | 7 (3.2) |
| Mornings, M-F (before 9am)* | 5 (2.3) |
| Mornings, M-F (before 9am), Evenings M-F (after 5pm) | 5 (2.3) |
| M-F (9am-5pm), Evenings M-F (after 5pm), Weekends (9am-5pm) | 4 (1.8) |
| Evenings M-F (after 5pm), Weekends (9am-5pm), Weekends (before 9am), Weekends (after 5pm) | 4 (1.8) |
| M-F (9am-5pm), Evenings M-F (after 5pm) | 4 (1.8) |
| Mornings, M-F (before 9am), Evenings M-F (after 5pm), Weekends (9am-5pm) | 3 (1.4) |
| Evenings M-F (after 5pm), Weekends (9am-5pm), Weekends (after 5pm) | 3 (1.4) |
| M-F (9am-5pm), Mornings, M-F (before 9am), Evenings M-F (after 5pm) | 2 (0.9) |
| Weekends (9am-5pm), Weekends (before 9am), Weekends (after 5pm) | 2 (0.9) |
| M-F (9am-5pm), Mornings, M-F (before 9am), Weekends (9am-5pm), Weekends (before 9am) | 2 (0.9) |
| Mornings, M-F (before 9am), Weekends (before 9am) | 2 (0.9) |
| M-F (9am-5pm), Mornings, M-F (before 9am), Weekends (9am-5pm) | 1 (0.5) |
| Mornings, M-F (before 9am), Weekends (9am-5pm) | 1 (0.5) |
| Weekends 9am-5pm, Weekends (before 9am) | 1 (0.5) |
| Evenings M-F (after 5pm), Weekends (9am-5pm), Weekends (before 9am) | 1 (0.5) |
| Mornings, M-F (before 9am), Evenings M-F (after 5pm), Weekends (after 5pm) | 1 (0.5) |
| Evenings M-F (after 5pm), Weekends (after 5pm) | 1 (0.5) |
| M-F (9am-5pm), Mornings, M-F (before 9am), Evenings M-F (after 5pm), Weekends (after 5pm) | 1 (0.5) |
| Weekends after 5pm* | 1 (0.5) |
| M-F (9am-5pm), Mornings, M-F (before 9am), Evenings M-F (after 5pm), Weekends (9am-5pm) | 1 (0.5) |
| Weekends (before 9am)* | 0 (0) |
| **Concerns about getting screened in a mobile screening unit – required question** |  |
| No concerns* | 160 (73.3) |
| Accurate results* | 8 (3.6) |
| Privacy* | 6 (2.7) |
| Cleanliness* | 5 (2.29) |
| Equipment, Cleanliness, Temperature, Privacy, Accurate results* | 4 (1.8) |
| Equipment, Cleanliness, Accurate results* | 4 (1.8) |
| Temperature, Privacy | 3 (1.4) |
| Cleanliness, Temperature, Privacy | 3 (1.4) |
| Equipment* | 3 (1.4) |
| Cleanliness, Privacy | 3 (1.4) |
| Equipment, Accurate results | 3 (1.4) |
| Equipment, Cleanliness, Privacy, Accurate results | 2 (0.9) |
| Cleanliness, Temperature | 2 (0.9) |
| Temperature* | 2 (0.9) |
| Privacy, Accurate results | 2 (0.9) |
| Cleanliness, Accurate results | 1 (0.5) |
| Equipment, Other | 1 (0.5) |
| Temperature, Accurate results | 1 (0.5) |
| Equipment, Cleanliness, Temperature, Privacy | 1 (0.5) |
| Equipment, Temperature, Privacy, Accurate results | 1 (0.5) |
| Other* | 1 (0.5) |
| Temperature, Privacy, Accurate results | 1 (0.5) |
| Cleanliness, Privacy, Accurate results | 1 (0.5) |

| **Survey Items** | **N (%)** |
| --- | --- |
| **Good time of year for a mobile lung cancer screening event – required question** |  |
| No preference* | 118 (54.1) |
| Fall* | 19 (8.7) |
| Spring, Fall, and Winter* | 17 (7.8) |
| Winter* | 17 (7.8) |
| Spring* | 10 (4.5) |
| Summer* | 8 (3.7) |
| Fall, and Winter* | 8 (3.7) |
| Spring, and Winter* | 6 (2.7) |
| Fall, Winter, Spring, and Summer* | 6 (2.7) |
| Spring, and Fall* | 3 (1.4) |
| Summer, and Fall* | 3 (1.4) |
| Spring and Summer | 1 (0.5) |
| Winter, Summer, and Fall | 1 (0.5) |
| Spring, Summer, and Fall | 1 (0.5) |
| **Easiest location for mobile bus screening – required question** |  |
| No preference* | 75 (34.4) |
| Neighborhood* | 16 (7.3) |
| Shopping mall/grocery store* | 10 (4.6) |
| Neighborhood, Shopping mall/grocery store* | 9 (4.1) |
| Neighborhood, Shopping mall/grocery store, and Church and other religious institutions* | 6 (2.7) |
| Neighborhood, health fair, and Church and other religious institutions* | 4 (1.8) |
| Downtown in my city* | 4 (1.8) |
| Neighborhood, Fire station or police station, Church and other religious institutions* | 4 (1.8) |
| Neighborhood, Fire station or police station, Shopping mall/grocery store* | 4 (1.8) |
| Neighborhood, Fire station or police station, Shopping mall/grocery store, health fair, school, Church and other religious institutions | 3 (1.4) |
| Neighborhood, Fire station or police station, Shopping mall/grocery store, school, post office, Church and other religious institutions | 3 (1.4) |
| Neighborhood, Fire station or police station, Shopping mall/grocery store, health fair, school, post office, Church and other religious institutions | 3 (1.4) |
| Neighborhood, Fire station or police station, Shopping mall/grocery store, post office | 3 (1.4) |
| Neighborhood, Fire station or police station, Shopping mall/grocery store, school | 3 (1.4) |
| Neighborhood, Fire station or police station, Shopping mall/grocery store, Church and other religious institutions | 2 (0.8) |
| Neighborhood, Shopping mall/grocery store, school, post office, Church and other religious institutions | 2 (0.8) |
| Fire station or police station, Shopping mall/grocery store, health fair, school | 2 (0.8) |
| Neighborhood, Fire station or police station, Shopping mall/grocery store, post office, Church and other religious institutions | 2 (0.8) |
| Shopping mall/grocery store, Church and other religious institutions | 2 (0.8) |
| Downtown in my city, Neighborhood, Shopping mall/grocery store, health fair | 2 (0.8) |
| Other* | 2 (0.8) |
| Fire station or police station* | 2 (0.8) |
| Neighborhood, Shopping mall/grocery store, health fair, school | 2 (0.8) |
| Neighborhood, health fair, other | 2 (0.8) |
| Fire station or police station, Shopping mall/grocery store, health fair, school, Church and other religious institutions | 2 (0.8) |
| Fire station or police station, Shopping mall/grocery store, school, Church and other religious institutions | 2 (0.8) |
| Fire station or police station, Church and other religious institutions | 2 (0.8) |
| Fire station or police station, school, Church and other religious institutions | 2 (0.8) |
| School* | 1 (0.5) |

| **Survey Items** | **N (%)** |
| --- | --- |
| Downtown in my city, Neighborhood, Fire station or police station, Shopping mall/grocery store, school, post office | 1 (0.5) |
| Shopping mall/grocery store, health fair, school, Church and other religious institutions | 1 (0.5) |
| Fire station or police station, school, health fair, Church and other religious institutions | 1 (0.5) |
| Neighborhood, school, Church and other religious institutions | 1 (0.5) |
| Downtown in my city, Neighborhood, Shopping mall/grocery store, school, other | 1 (0.5) |
| Fire station or police station, health fair | 1 (0.5) |
| health fair, school, Church and other religious institutions | 1 (0.5) |
| Shopping mall/grocery store, school, Church and other religious institutions | 1 (0.5) |
| Neighborhood, Shopping mall/grocery store, health fair, school | 1 (0.5) |
| Downtown in my city, Neighborhood, Fire station or police station, Shopping mall/grocery store, post office | 1 (0.5) |
| Shopping mall/grocery store, health fair | 1 (0.5) |
| Neighborhood, health fair, school, other | 1 (0.5) |
| Shopping mall/grocery store, other | 1 (0.5) |
| Neighborhood, Fire station or police station, Shopping mall/grocery store, school, post office | 1 (0.5) |
| Shopping mall/grocery store, health fair, school | 1 (0.5) |
| health fair, other | 1 (0.5) |
| Neighborhood, school, post office, Church and other religious institutions | 1 (0.5) |
| Downtown in my city, Neighborhood, Shopping mall/grocery store, post office | 1 (0.5) |
| Downtown in my city, Fire station or police station, health fair | 1 (0.5) |
| Church and other religious institutions* | 1 (0.5) |
| Downtown in my city, Neighborhood | 1 (0.5) |
| Fire station or police station, school, post office, Church and other religious institutions | 1 (0.5) |
| Downtown in my city, Shopping mall/grocery store | 1 (0.5) |
| Neighborhood, Shopping mall/grocery store, health fair, Church and other religious institutions | 1 (0.5) |
| Downtown in my city, Neighborhood, Fire station or police station, Shopping mall/grocery store | 1 (0.5) |
| Neighborhood, Fire station or police station, health fair, school | 1 (0.5) |
| Fire station or police station, Shopping mall/grocery store, Church and other religious institutions | 1 (0.5) |
| Neighborhood, Fire station or police station, Shopping mall/grocery store, health fair, school | 1 (0.5) |
| Neighborhood, Shopping mall/grocery store, post office | 1 (0.5) |
| Downtown in my city, Neighborhood, Fire station or police station | 1 (0.5) |
| Neighborhood, Shopping mall/grocery store, school | 1 (0.5) |
| Neighborhood, health fair | 1 (0.5) |
| Neighborhood, school | 1 (0.5) |
| Neighborhood, Shopping mall/grocery store, school, post office, other | 1 (0.5) |
| Downtown in my city, Neighborhood, Shopping mall/grocery store, health fair, Church and other religious institutions | 1 (0.5) |
| Fire station or police station, health fair, Church and other religious institutions | 1 (0.5) |
| Downtown in my city, Fire station or police station, health fair, post office | 1 (0.5) |
| Neighborhood, Fire station or police station, Shopping mall/grocery store, health fair | 1 (0.5) |
| Neighborhood, Fire station or police station, Shopping mall/grocery store, school, Church and other religious institutions | 1 (0.5) |
| Fire station or police station, Shopping mall/grocery store, health fair | 1 (0.5) |
| Downtown in my city, Neighborhood, Fire station or police station, school, Church and other religious institutions | 1 (0.5) |
| Shopping mall/grocery store, school, post office Church and other religious institutions | 1 (0.5) |
| Health fair* | 0 (0) |
| Post office* | 0 (0) |
| **Previous experiences with cancer screenings?** |  |
| Breast, Colon, Skin, and Cervical* | 35 (16) |
| Breast, Colon, Lung, Skin, and Cervical* | 17 (7.7) |
| Breast* | 15 (6.8) |
| Breast, Colon, and Skin* | 12 (5.5) |

| **Survey Items** | **N (%)** |
| --- | --- |
| Colon, Prostate, and Skin* | 11 (5.0) |
| Breast, Colon, and Cervical* | 9 (4.1) |
| Breast, Skin, and Cervical* | 8 (3.7) |
| Breast, and Colon* | 8 (3.7) |
| Breast, Colon, Lung, and Skin | 6 (2.7) |
| Breast, and Lung | 6 (2.7) |
| Breast, Lung, and Skin | 6 (2.7) |
| Breast, and Other | 6 (2.7) |
| Prostate* | 5 (2.3) |
| Breast, and Skin | 5 (2.3) |
| Lung* | 4 (1.8) |
| Colon* | 4 (1.8) |
| Colon, Lung, Prostate, and Skin | 4 (1.8) |
| Breast, and Cervical | 4 (1.8) |
| Prostate, and Skin | 4 (1.8) |
| Colon, and Skin | 3 (1.4) |
| Missing* | 3 (1.4) |
| Breast, Lung, and Cervical | 3 (1.4) |
| Other* | 3 (1.4) |
| No* | 3 (1.4) |
| Breast, Colon, Lung, and Cervical | 3 (1.4) |
| Breast, Colon, and Lung | 3 (1.4) |
| Lung, and Prostate | 2 (0.9) |
| Breast, Colon, Skin, and Other | 2 (0.9) |
| Breast, Colon, Skin, and Other | 2 (0.9) |
| Breast, Skin, and Other | 2 (0.9) |
| Skin* | 2 (0.9) |
| Breast, Lung, Skin, and Cervical | 2 (0.9) |
| Colon, and Prostate | 2 (0.9) |
| Colon, Prostate, Skin, and Other | 1 (0.5) |
| Breast, Colon, Lung, Skin, Cervical, and Other | 1 (0.5) |
| Breast, Colon, Lung, Skin, and Other | 1 (0.5) |
| Colon, Lung, Skin, and Other | 1 (0.5) |
| Colon, Lung, Prostate, Skin, and Other | 1 (0.5) |
| Colon, and Lung | 1 (0.5) |
| Lung, Prostate, and Skin | 1 (0.5) |
| Breast, Colon, and Other | 1 (0.5) |
| Colon, Lung, and Prostate | 1 (0.5) |
| Breast, Lung, Skin, and Other | 1 (0.5) |
| Colon, Lung, and Skin | 1 (0.5) |
| Skin, and Other | 1 (0.5) |
| Colon, Skin, and Cervical | 1 (0.5) |
| Lung, Other | 1 (0.5) |
| Cervical* | 0(0) |
| **If you wanted more information about mobile lung cancer screening, how would you want to receive this information?** |  |
| Receiving a notification in the mail* | 26 (11.9) |
| Talking to your primary doctor* | 20 (9.2) |
| Talking to your primary doctor, Receiving a message through your electronic health record, and Receiving a notification in the mail* | 15 (6.9) |
| Receiving a message through your electronic health record* | 14 (6.4) |
| Talking to your primary doctor, and Receiving a message through your electronic health record* | 9 (4.1) |
| Receiving a message through your electronic health record, and Receiving a notification in the mail* | 8 (3.7) |
| Talking to your primary doctor, and Receiving a notification in the mail* | 7 (3.2) |

| **Survey Items** | **N (%)** |
| --- | --- |
| Talking to your primary doctor, Receiving a phone call, Receiving a message through your electronic health record, and Receiving a notification in the mail* | 7 (3.2) |
| Receiving a phone call, Receiving a message through your electronic health record, and Receiving a notification in the mail* | 6 (2.7) |
| Talking to your primary doctor, Receiving a phone call, and Receiving a message through your electronic health record* | 6 (2.7) |
| Talking to your primary doctor, Receiving a notification in the mail, and Watching a local news segment or television special about this | 4 (1.7) |
| Receiving a phone call* | 4 (1.7) |
| Other* | 4 (1.7) |
| Talking to your primary doctor, Receiving a message through your electronic health record, Receiving a notification in the mail, and Watching a local news segment or television special about this | 3 (1.3) |
| Talking to your primary doctor, and Other | 3 (1.3) |
| Through social media (for example, Facebook, Twitter, Instagram), and Receiving a notification in the mail | 3 (1.3) |
| Talking to your primary doctor, and Reading about this in a magazine/newspaper you subscribe to | 3 (1.3) |
| Through social media (for example, Facebook, Twitter, Instagram), Talking to your primary doctor, Watching a local news segment or television special about this, and Reading about this in a magazine/newspaper you subscribe to | 3 (1.3) |
| Receiving a notification in the mail, and Other | 3 (1.3) |
| Talking to your primary doctor, and Receiving a phone call | 3 (1.3) |
| Talking to your primary doctor, Receiving a notification in the mail, and Other | 3 (1.3) |
| Through social media (for example, Facebook, Twitter, Instagram), and Talking to your primary doctor | 3 (1.3) |
| Through social media (for example, Facebook, Twitter, Instagram), Talking to your primary doctor, Receiving a message through your electronic health record, and Watching a local news segment or television special about this | 2 (0.9) |
| Receiving a phone call, Receiving a message through your electronic health record, Receiving a notification in the mail, and Other | 2 (0.9) |
| Through social media (for example, Facebook, Twitter, Instagram), Talking to your primary doctor, and Receiving a message through your electronic health record | 2 (0.9) |
| Receiving a phone call, and Receiving a message through your electronic health record | 2 (0.9) |
| Through social media (for example, Facebook, Twitter, Instagram), and Watching a local news segment or television special about this | 2 (0.9) |
| Talking to your primary doctor, Receiving a message through your electronic health record, Receiving a notification in the mail, and Other | 2 (0.9) |
| Through social media (for example, Facebook, Twitter, Instagram), Receiving a notification in the mail, and Watching a local news segment or television special about this | 2 (0.9) |
| Through social media (for example, Facebook, Twitter, Instagram), Talking to your primary doctor, Receiving a message through your electronic health record, Receiving a notification in the mail, Watching a local news segment or television special about this, and Reading about this in a magazine/newspaper you subscribe to | 2 (0.9) |
| Through social media (for example, Facebook, Twitter, Instagram), Talking to your primary doctor, Receiving a phone call, Receiving a message through your electronic health record, Receiving a notification in the mail, Watching a local news segment or television special about this, and Reading about this in a magazine/newspaper you subscribe to | 2 (0.9) |
| Missing* | 2 (0.9) |
| Through social media (for example, Facebook, Twitter, Instagram)* | 2 (0.9) |
| Through social media (for example, Facebook, Twitter, Instagram), Talking to your primary doctor, and Receiving a notification in the mail | 2 (0.9) |
| Receiving a notification in the mail, Watching a local news segment or television special about this, and Reading about this in a magazine/newspaper you subscribe to | 2 (0.9) |
| Receiving a notification in the mail, Watching a local news segment or television special about this, Reading about this in a magazine/newspaper you subscribe to, and Other | 1 (0.5) |
| Receiving a phone call, and Receiving a notification in the mail | 1 (0.5) |

| **Survey Items** | **N (%)** |
| --- | --- |
| Talking to your primary doctor, Receiving a message through your electronic health record, Receiving a notification in the mail, Watching a local news segment or television special about this, and Other | 1 (0.5) |
| Receiving a phone call, and Other | 1 (0.5) |
| Through social media (for example, Facebook, Twitter, Instagram), Talking to your primary doctor, and Receiving a phone call | 1 (0.5) |
| Talking to your primary doctor, Receiving a notification in the mail, and Reading about this in a magazine/newspaper you subscribe to | 1 (0.5) |
| Receiving a phone call, Receiving a notification in the mail, and Watching a local news segment or television special about this | 1 (0.5) |
| Receiving a phone call, Receiving a notification in the mail, Watching a local news segment or television special about this, and Reading about this in a magazine/newspaper you subscribe to | 1 (0.5) |
| Through social media (for example, Facebook, Twitter, Instagram), Talking to your primary doctor, and Watching a local news segment or television special about this | 1 (0.5) |
| Through social media (for example, Facebook, Twitter, Instagram), Talking to your primary doctor, Receiving a message through your electronic health record, and Other | 1 (0.5) |
| Talking to your primary doctor, Receiving a phone call, Receiving a message through your electronic health record, Receiving a notification in the mail, and Watching a local news segment or television special about this | 1 (0.5) |
| Through social media (for example, Facebook, Twitter, Instagram), Receiving a phone call, and Receiving a notification in the mail | 1 (0.5) |
| Receiving a message through your electronic health record, Receiving a notification in the mail, Watching a local news segment or television special about this, and Other | 1 (0.5) |
| Through social media (for example, Facebook, Twitter, Instagram), Talking to your primary doctor, Receiving a message through your electronic health record, Watching a local news segment or television special about this, and Other | 1 (0.5) |
| Through social media (for example, Facebook, Twitter, Instagram), Talking to your primary doctor, Receiving a message through your electronic health record, and Reading about this in a magazine/newspaper you subscribe to | 1 (0.5) |
| Receiving a notification in the mail, Watching a local news segment or television special about this and Other | 1 (0.5) |
| Receiving a phone call, Receiving a message through your electronic health record, Receiving a notification in the mail, Watching a local news segment or television special about this, and Reading about this in a magazine/newspaper you subscribe to | 1 (0.5) |
| Reading about this in a magazine/newspaper you subscribe to* | 1 (0.5) |
| Talking to your primary doctor, Receiving a phone call, Receiving a message through your electronic health record, and Other | 1 (0.5) |
| Talking to your primary doctor, Receiving a message through your electronic health record, and Other | 1 (0.5) |
| Talking to your primary doctor, Receiving a notification in the mail, Watching a local news segment or television special about this, and Other | 1 (0.5) |
| Receiving a message through your electronic health record, and Other | 1 (0.5) |
| Through social media (for example, Facebook, Twitter, Instagram), Receiving a message through your electronic health record, Receiving a notification in the mail, Watching a local news segment or television special about this, and Reading about this in a magazine/newspaper you subscribe to | 1 (0.5) |
| Watching a local news segment or television special about this* | 1 (0.5) |
| Through social media (for example, Facebook, Twitter, Instagram), Talking to your primary doctor, Receiving a phone call, and Watching a local news segment or television special about this | 1 (0.5) |
| Through social media (for example, Facebook, Twitter, Instagram), Talking to your primary doctor, Receiving a notification in the mail, Watching a local news segment or television special about this, and Reading about this in a magazine/newspaper you subscribe to | 1 (0.5) |
| Through social media (for example, Facebook, Twitter, Instagram), Talking to your primary doctor, Receiving a message through your electronic health record, and Receiving a notification in the mail | 1 (0.5) |
| Through social media (for example, Facebook, Twitter, Instagram), Receiving a message through your electronic health record, Watching a local news segment or television special about this, and Reading about this in a magazine/newspaper you subscribe to | 1 (0.5) |

| **Survey Items** | **N (%)** |
| --- | --- |
| Receiving a message through your electronic health record, Watching a local news segment or television special about this, and Reading about this in a magazine/newspaper you subscribe to | 1 (0.5) |
| Talking to your primary doctor, Receiving a message through your electronic health record, Watching a local news segment or television special about this, and Reading about this in a magazine/newspaper you subscribe to | 1 (0.5) |
| Through social media (for example, Facebook, Twitter, Instagram), and Receiving a message through your electronic health record | 1 (0.5) |
| Talking to your primary doctor, and Watching a local news segment or television special about this | 1 (0.5) |
| HER, Receiving a notification in the mail, and Watching a local news segment or television special about this | 1 (0.5) |
| Talking to your primary doctor, Receiving a message through your electronic health record, and Watching a local news segment or television special about this | 1 (0.5) |
| Through social media (for example, Facebook, Twitter, Instagram), Talking to your primary doctor, and Other | 1 (0.5) |
| **If you needed a follow-up appointment after your lung cancer screening, where would you be willing to have it?** |  |
| Hospital/Clinic, Mobile Screening Unit, and Virtual* | 132 (60.5) |
| Hospital/clinic Only* | 49 (22.4) |
| Virtual Only* | 2 (0.9) |
| Mobile Screening Unit Only* | 0 (0) |
| Hospital/Clinic, and Mobile Screening Unit* | 12 (5.5) |
| Hospital/Clinic and Virtual* | 9 (4.1) |
| Other* | 9 (4.1) |
| Mobile Screening Unit, and Other* | 1 (0.5) |
| Virtual and Other* | 1 (0.5) |
| Hospital/clinic, Virtual, and Other* | 1 (0.5) |
| All the above, and Other* | 1 (0.5) |
| Hospital/Clinic, Mobile Screening Unit and Other* | 1 (0.5) |

\*Responses also reported in Table 2

